# Using computable knowledge mined from the literature to elucidate confounders for EHR-based pharmacovigilance

**DOI:** 10.1101/2020.07.08.20113035

**Authors:** Scott A. Malec, Peng Wei, Elmer V. Bernstam, Richard D. Boyce, Trevor Cohen

## Abstract

**Introduction:** Drug safety research asks causal questions but relies on observational data. Confounding bias threatens the reliability of studies using such data. The successful control of confounding requires knowledge of variables called confounders affecting both the exposure and outcome of interest. Causal knowledge of dynamic biological systems is complex and challenging. Fortunately, computable knowledge mined from the literature may hold clues about confounders. In this paper, we tested the hypothesis that incorporating literature-derived confounders can improve causal inference from observational data.

**Methods:** We introduce two methods (semantic vector-based and string-based confounder search) that query literature-derived information for confounder candidates to control, using SemMedDB, a database of computable knowledge mined from the biomedical literature. These methods search SemMedDB for confounders by applying semantic constraint search for indications treated by the drug (exposure), that are also known to cause the adverse event (outcome). We then include the literature-derived confounder candidates in statistical and causal models derived from free-text clinical notes. For evaluation, we use a reference dataset widely used in drug safety containing labeled pairwise relationships between drugs and adverse events and attempt to rediscover these relationships from a corpus of 2.2M NLP-processed free-text clinical notes. We employ standard adjustment and causal inference procedures to predict and estimate causal effects by informing the models with varying numbers of literature-derived confounders and instantiating the exposure, outcome, and confounder variables in the models with dichotomous EHR-derived data. Finally, we compare the results from applying these procedures with naive measures of association (*χ*^2^ and reporting odds ratio) and with each other.

**Results and Conclusions:** We found semantic vector-based search to be superior to string-based search at reducing confounding bias. However, the effect of including more rather than fewer literature-derived confounders was inconclusive. We recommend using targeted learning estimation methods that can address treatment-confounder feedback, where confounders that also behave as intermediate variables, and engaging subject-matter experts to adjudicate the handling of problematic confounders.

**Highlights:**

- Drug safety research asks causal questions but must rely on observational data
- We explore searching literature-derived computable knowledge to elucidate confounders
- To identify confounders, we search for common causes relative to a drug and an adverse event
- We test search, modeling, and inference procedures on EHR data to detect genuine adverse events
- Semantic vector-based search performed better overall than string-based confounder search.

**Graphical Abstract:** 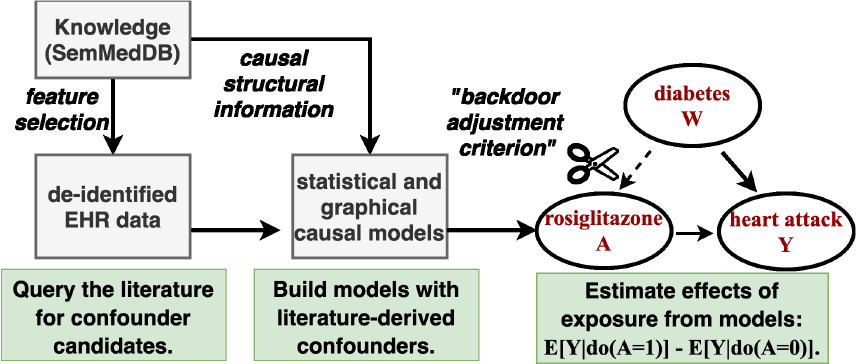

## 1. Introduction

This paper introduces a framework for automating causal inference from observational data by exploiting computable knowledge mined from the literature. Observational data, or data collected in non-randomized settings, contain a wealth of information for biomedical research. Such data are vital in settings where randomized controlled trials are infeasible, such as is the case with drug safety research.

A primary goal of drug safety (or pharmacovigilance) research is to prioritize associations empirically to the extent that drug / adverse event associations may be causal or stochastically determinative in nature. Associations that are more likely to be genuinely causative should be prioritized for review. Unfortunately, confounding, endemic to such data, can induce misleading, non-causal associations. While the research question defines the exposure and outcome variables, the decision of which covariates to adjust for falls to the investigator. Classical criteria mandate for the inclusion of all covariates correlating significantly with the exposure and the outcome [1]. Unfortunately, such an approach can introduce covariates that amplify bias rather than reduce it [2, 3, 4].

Recently, to address the issue of problematic covariates, researchers have enumerated criteria for identifying adjustment sets emphasizing the role of causal knowledge for identifying covariates to reduce bias [5]. Unfortunately, since such expertise cannot scale to all available human knowledge, it is infeasible to rely solely on human experts. Consequently, the problem of how to access causal knowledge has been noted as an open research question [6]. Fortunately, knowledge resources and methods exist that could be useful for guiding the selection of confounders. The Semantic MEDLINE database, or SemMedDB, is one such resource [7]. The information in SemMedDB consists of pairs of biomedical entities, or concepts, connected by normalized predicates, e.g., “aspirin TREATS headache.” We introduce and test methodological variants that combine computable knowledge with observational data. Our methods query literature-derived computable knowledge to identify confounder candidates for incorporation into statistical and graphical causal models. The idea is to use existing knowledge from previous discoveries to catalyze causal inference and discovery from observational data. We then use these models to perform statistical and causal inference from data extracted from a corpus of EHR-derived free-text clinical narratives.

The purpose of this research is to investigate the extent to which computable knowledge may be useful for informing causal inference. To that end, we introduce and test two methods for accessing background knowledge to help reduce confounding bias: string-based and semantic vector-based search. These methods are qualitatively distinct in how they store, represent, and retrieve information. We also explore whether or not knowledge representation affects performance, given how concepts are prioritized by the confounder search methods in search results. We also ask how varying the amount of literature-derived information affects bias reduction. Finally, we compare using effect estimates from literature-informed causal models with traditional logistic regression adjustment procedures.

## 2. Background

With nearly half of the US population having been prescribed a prescription drug in any given month, the vast number of drug exposures drives the high prevalence of adverse events [8]. The annual financial cost of adverse event-related morbidity in the United States was estimated at 528.4 billion in 2016 alone [9], while another study noted that 16.88% of hospitalized patients experience an adverse drug reaction [10]. An adverse drug reaction is defined as an “appreciably harmful or unpleasant reaction, resulting from an intervention related to the use of a medical product” [11], and is distinguished from other adverse events by the demonstration of a causal link to a drug. The discipline that helps adjudicate causal links between drugs and harmful side-effects is known as pharmacovigilance. Pharmacovigilance encompasses the development of procedures for collecting, summarizing, monitoring, detecting, and reviewing associations between drug exposures and health outcomes in general and adverse events in particular [12]. Causal links are established by the gradual accretion of evidence from observational data and by the relative strength of mechanistic explanations justifying biological plausibility [13].

In medicine, randomized controlled trials (RCTs) are widely considered to be the gold standard for establishing causal links [14]. However, in research areas such as pharmacovigilance, the relevance of RCTs is limited most pertinently by ethical considerations but further by the size, cost, and short duration of such studies [14, 15]. Accordingly, to prioritize drug safety signals for review after regulatory approval of new drugs, regulatory bodies such as the Food and Drug Administration in the United States and the European Medicines Agency in the European Union must rely mainly on the quality of the inferences drawn from empirical data from non-randomized sources. The traditional primary data source for pharmacovigilance research has been spontaneous reporting systems such as the U.S. Food and Drug Administration (FDA) Adverse Event Reporting System (FAERS) [16, 17]. Spontaneous reporting systems’ data are aggregated from input from patients, clinicians, and pharmaceutical companies. Unfortunately, these data present critical deficiencies, including lack of context, missing data, and no population-level denominator to estimate the prevalence of any association [18, 19]. Consequently, there is a pressing need to advance methods that can more reliably detect adverse drug reactions from observational data.

To address the shortcomings of spontaneous reporting systems, researchers have turned to other sources of empirical evidence such as social media [20, 21], claims [22, 23], and EHR data [24, 25, 26, 27], the focus of the present study. The FDA’s ongoing Sentinel Initiative facilitates the federated search of structured data from EHR across the United States [28]. However, structured data may only provide an incomplete picture. Data embedded in unstructured free-text clinical narratives in EHR systems contain a wealth of contextual information concerning routine clinical practice often absent from the fixed content fields in structured data [29]. The embedded contextual information may include “temporal relations, severity and degree modifiers, causal connections, clinical explanations, and rationale” [30].

The reliability of analytic conclusions from such data is highly sensitive to the quality of the assumptions used to analyze them [31, 32], including assumptions concerning how to adjust for sources of systematic bias. The focus of the present study is on confounding bias, though other forms of systematic bias exist [3].

In randomized experiments, confounding control is built-in by design. However, in studies using observational data, variables of interest are subject to sources of influence outside investigator control. In this setting, the investigator must identify variables for which to control by analysis.

A confounder is a type of variable that is relative to the hypothesis under study that influences both the likelihood of the exposure and the outcome (Figure 1) [33]. Confounding bias is induced when both the exposure and outcome of interest share a common cause that is not controlled analytically.

**Figure 1:**
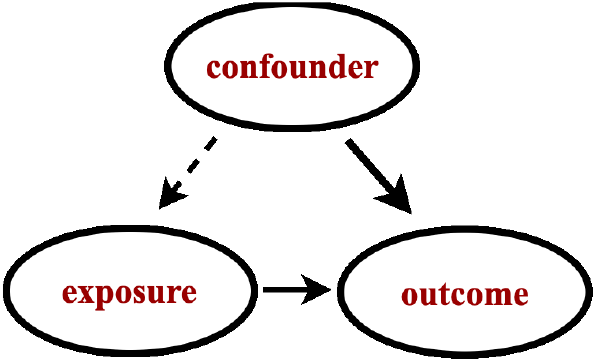
Illustration of confounding. If both the exposure and outcome variables are effects of the same cause. That common cause is referred to as a confounder.

If it were possible to infer which biomedical entities, or concepts, might also refer to particular variables for which to control, i.e., confounders, it would be possible to facilitate automated discovery from non-randomized settings from observational data [34].

Clearly, substantive, extra-statistical *a priori* subject-specific knowledge is critical for reliable causal inference [5, 32]. Since it has not been clear how to access contextually relevant causal knowledge automatically, many approaches for inferring causality have been developed to evade the requirement of subject-specific knowledge. For example, the approach described in [35] employs meta-analytic techniques by combining different data sources (EHR data with FAERS) with the idea that such combination cancels out bias from individual sources of data. Other approaches impute a pseudo-variable to absorb residual confounding between measured variables [36, 37, 38].

However, our ongoing research’s focus assumes the existence of computable knowledge mined from the literature to help select covariates to facilitate causal inference. Computable knowledge mined from the biomedical literature in the form of machine readable concept-relation-concept semantic predications could be useful for identifying relevant biomedical concepts such as confounders given a drug exposure and a health outcome of interest. This paper’s primary contribution is the description of how standard epidemiological definitions can be used to map concepts in literature-derived computable knowledge to observed, measured variables in free-text clinical narrative data derived from EHR relevant for controlling confounding bias.

### 2.1. Components of a causal inference toolkit

In the next section, we introduce background material vital to understanding our methods in terms of the essential components and procedures. Figure 2 illustrates our toolkit’s components for leveraging background knowledge from the literature to catalyze causal inference.

**Figure 2:**
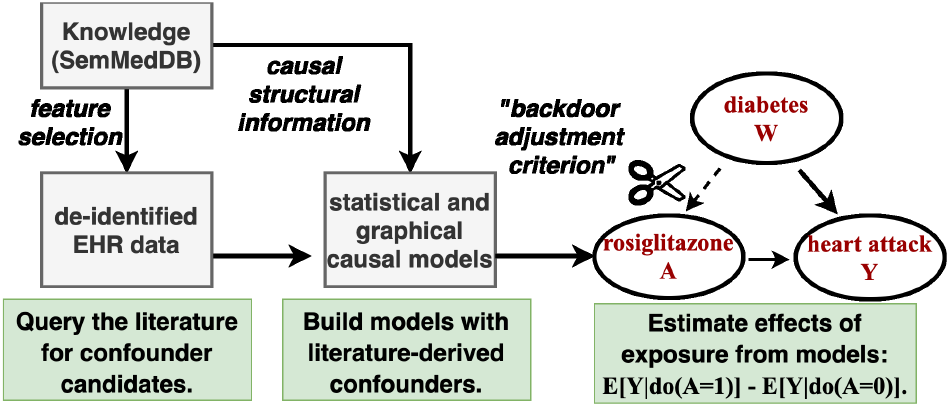
Schema depicting our literature-informed causal modeling toolkit.

#### 2.1.1. Causal inference

The field of causal inference concerns itself with the development of methods for estimating causal parameters of interest. Note that in the causal inference literature, the exposure is often referred to as the “treatment,” and we shall use both terms interchangeably (also, treated and untreated can stand in for exposed and unexposed).

A common goal of causal inference is to estimate the average treatment effect (ATE). The ATE represents an expectation (or population-level average, or mean) quantifying the extent to which an intervention (such as a drug exposure) would affect an outcome of interest (such as an adverse event), given two cohorts (one exposed and the other unexposed) with similar characteristics. Causal inference is achieved by calculating the mean difference in potential outcomes across exposed and unexposed subgroups with similar pre-exposure characteristics, or confounders. The *do*(.) operator first introduced by Pearl was invented to complement conventional mathematical notation to denote such an operation [34].

For a binary (non-dose-dependent) treatment **A** with effect **Y**, ATE can be estimated as a contrast between two levels: the exposed and unexposed groups given observed confounders **W**. The lowercase *a* denotes the variable being fixed to set value, where **a** = 1 for exposed and **a** = 0 for unexposed. (Henceforth, we shall refer to drug exposures and treatments interchangeably.) The ATE (henceforth denoted Δ) is expressed as the following equation:

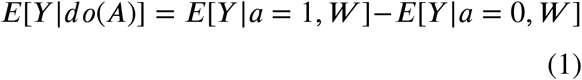

This equation defines the adjusted treatment effect, and adjusts for confounders **W**. However, in order for Equation 1 to estimate causal effects reliably, these confounders must first be identified. Consequently, the next important task is to determine the features or variables **W** for which to adjust.

To identify confounders, causal theory has introduced the notion of backdoor paths or paths that point into the exposure, resulting in a mixture of both the confounder’s and the exposure’s effects. The backdoor-criterion stipulates that causal inference is possible if a set of covariates **W** can be found that block all “backdoor” paths from **A** to **Y**. If so, then bias from confounding can be reduced or eliminated [34].

In the next section, we discuss computable knowledge resources and the methods that can reason over large volumes of computable knowledge extracted from the published biomedical literature to provide convenient access to identify contextually relevant knowledge.

#### 2.1.2. Literature-based discovery

The objective of literature-based discovery research (henceforth, LBD) is to reveal meaningful but implicit connections between biomedical entities of interest in the published literature [39, 40, 41]. The late Don Swanson pioneered LBD in his seminal work, discovering the potential of fish oil to treat Raynaud’s syndrome [42], an example we will return to below. Much early work in LBD focused on investigating the strength of association between concepts and exploiting insights from information retrieval into concept co-occurrence patterns. However, concept co-occurrence alone can produce more hypotheses than it is possible to review.

To further constrain the results returned by LBD systems, researchers have developed ways to exploit information concerning the nature of the relationships between biomedical concepts. Revisiting Swanson’s original example with fish oil, researchers noticed that it was useful to pay attention not only to concepts themselves but to information concerning *relationships between concepts* [43, 44]. For example, certain drug concepts are known to treat diseases, and so the drug and the disease are related through a TREATS relationship. Other drug exposures may be known as causal agents of the adverse events in the published literature, and thus be related via the CAUSES predicate, i.e., Vioxx CAUSES acute_myocardial_infarction.

To demonstrate how such knowledge can be useful, consider the example of Raynaud’s disease, from Don Swanson’s work [42]. Raynaud’s is a circulatory disorder manifesting in skin discoloration affecting the extremities. Blood viscosity (loosely defined as “thickness” or “stickiness”) is implicated as a mechanism in Raynaud’s, [42, 45], with **increasing** viscosity in cold conditions thought to impede circulation to the peripheral extremities causing them to appear white or blue, the primary symptom of Raynaud’s. Swanson noticed that fish oil could have the effect of **decreasing** blood viscosity, thus leading to his therapeutic hypothesis that fish oil can treat Raynaud’s by countering the mechanisms of Raynaud’s. Extrapolating from Swanson’s example, researchers paid attention to how concepts were related to each other (e.g., A increases B, B decreases C in the example above), and were able to manually extrapolate useful patterns, called discovery patterns, that could generate biologically plausible hypotheses for novel therapies [46, 43]. Discovery patterns define semantic constraints for identifying concepts related to each other in particular ways [46].

We conjecture that standard epidemiological definitions for confounding may be translated into discovery patterns for identifying concepts describing confounders for which to adjust to enhance inference from observational data by reducing confounding bias.

#### 2.1.3. SemMedDB - a causal knowledge resource

SemMedDB is a knowledge database deployed extensively in biomedical research and developed at the US National Library of Medicine. The knowledge contained in SemMedDB consists of subject-predicate-object triples (or predications) extracted from titles and abstracts in MEDLINE [47] using the SemRep biomedical NLP system [47, 48, 49]. SemRep can be thought of as a machine reading utility for transforming biomedical literature into computable knowledge. Employing a rule-based syntactic parser enriched with domain knowledge, SemRep first uses the high precision MetaMap [50] (e.g.,estimated at 83% in [48]) biomedical concept tagger to recognize biomedical entities (or concepts) in the Unified Medical Language System (UMLS). The UMLS is a compendium of codes and terms for representing concepts across many health and biomedical domains used to enable semantic processing and translation across terminologies [51].

Next, SemRep categorizes how the recognized concepts are associated given a fixed set of normalized, pre-specified predicates (with thirty core predicate types) corresponding to relations of biomedical interest, e.g., CAUSES, PREDISPOSES, TREATS, PREVENTS, STIMULATES, INHIBITS, AFFECTS [51, 48]. For example, the predication “ibuprofen TREATS inflammation_disorder” was extracted by SemRep from the source text: “Ibuprofen has gained widespread acceptance for the treatment of rheumatoid arthritis and other inflammatory disorders.” In the next subsection, we describe a knowledge representation scheme for accessing computable knowledge extracted from the literature.

#### 2.1.4. Predication-based Semantic Indexing (PSI)

Predication-based semantic indexing, or PSI, defines a scheme for encoding and performing approximate inference over large volumes of computable knowledge [52]. The basic premise of distributional semantics is that terms that appear in similar contexts tend to have similar meanings [53].

By encoding the contexts in which terms appear, methods of distributional semantics provide a natural way to extrapolate semantics automatically from a corpus. PSI’s approach to distributional semantics derives from the Random Indexing (RI) paradigm, wherein a semantic vector for each term is created as the (possibly weighted) sum of randomly instantiated vectors - which we will refer to as *elemental vectors* as they are not altered during training - representing the contexts in which it occurs. [54, 55]. To adapt the RI approach for encoding concept-relation-concept triples extracted from the literature by SemRep, PSI adopts an approach that is characteristic of a class of representational frameworks collectively known as vector-symbolic architectures [56, 57, 58], or VSAs. VSAs were developed in response to a debate (with one side forcefully articulated in [59]) concerning the ability to represent hierarchical structures in connectionist models of cognition.

In RI, elemental vectors of high dimensionality (of dimensionality ≥. 1000), with a small number of non-zero values (≥ 10) which are set to either +1 or −1 at random, are generated for each context (where a context might represent a document or the presence of some other term in proximity to the term to be represented). The resulting vectors have a high probability of being approximately orthogonal, ensuring that each context has a distinct pattern that acts as a fingerprint for it. Alternatively, and as is the case with the current research, high-dimensional binary vectors (of dimensionality ≥ 10000 *bits*) can be employed as a unit of representation. In this case, vectors are initialized with an equal number of 0s and 1s, assigned randomly. While these vectors are not sparse in the “mostly zero” sense, they retain the desirable property of approximate orthogonality [60], with orthogonality defined as a hamming distance of half the vector dimensionality.

VSAs provide an additional mechanism for encoding structured information by using what is known as a “binding operator”, a nomenclature that suggests its application as a means to bind variables to values within the connectionist representational paradigm. The binding operator is an invertible operator that combines two vector representations *a* and *b*, to form a third *c* that is dissimilar to its component vectors. As this operation is invertible, *a* can be recovered from *c* using *b*, and vice-versa. Thus, through the invertibility property of the binding operation, a value can be recovered from a variable to which it is bound, thereby facilitating retrieval from the encoded computable knowledge.

PSI is implemented in the open-source and publicly available `Semantic Vectors` package written in Java [61], and in the context of SemMedDB is applied to concepts which are defined by normalized biomedical entities in the UMLS hierarchy, rather than the original terms as encountered in the biomedical source text.

The resulting models have been applied to a range of biomedical problems (as reviewed in [62]). PSI accepts semantic predications as input and transforms that input into a searchable vector space that may then be used as a search engine to retrieve confounders. Next, we describe how PSI spaces are built and introduce semantic vector algebra, a convenient shorthand to describe the training processes of PSI spaces and to compose useful queries over the resulting PSI spaces.

##### Training a PSI space

The process for training PSI begins by making an elemental vector, denoted *E*(.), for each unique concept and each unique relation, in the corpus. PSI constructs semantic vectors, denoted *S*(.), by superposing, denoted +=, the bound product of the elemental vectors of each relation and concept with which it co-occurs. The binding operation, denoted ⊗ is invertible, and its inverse, called ‘release,’ is denoted ⊘. PSI encodes the semantic predication “aspirin TREATS headaches” like so:

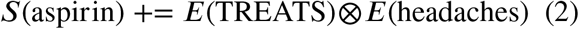

Note that for directional predications, the inverse predicate is also encoded. For a predicate like “TREATS,” the meaning would be “TREATED BY,” here denoted TREATS_INV_, thus:

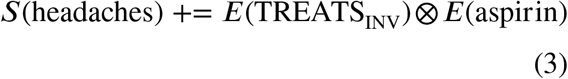

However, no such representation is made for predicates lacking “direction” such as COEXISTS_WITH and ASSOCIATED_WITH. These predicates are their own inverse.

After a PSI model has been trained, the resulting PSI space may be searched. As we explain below, the process of querying the PSI space reverses the operations used in the training procedure.

##### Querying a PSI space

PSI can perform approximate inference over the knowledge it has encoded after a semantic space has been constructed from the semantic predications. The `Semantic Vectors` package [63, 64] implements a VSA-based query language that facilitates queries to the resulting vector space, which underlies the `EpiphaNet` system for literature-based discovery [65].

The `Semantic Vectors` query language provides a calculus for using vector algebra to perform logical inference over the computable knowledge contained within the vector space, enabling discovery pattern-based search. It provides a functional implementation of the binding and superposition operators used in VSAs. However, in this implementation it is important to note that binding and release are equivalent (⊗ == ⊘), and as such are *both* represented with the symbol ⊗ (this follows from the use of pairwise XOR as a self-invertible binding operator in binary vector based PSI implementations). In this calculus, *P* (.) denotes an elemental predicate vector (while, as mentioned earlier, *S*(.) and *E*(.) denote a semantic vector or elemental vectors for a concept, respectively). We demonstrate the application of this syntax using an example composite PSI query for searching for concepts treated by the anti-diabetic drug rosiglitazone, that also causes heart attacks.

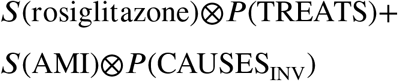

In the above PSI query, the syntax denotes a binding (or release - ⊗) operation between the semantic vector of rosiglitazone with the predicate vector (of the predicate TREATS). The resulting bound product S(rosiglitazone) ⊗ P(TREATS) is a vector that we would anticipate being similar to the elemental (random index) vectors for the terms representing such entities. In the next component of the query, the semantic vector for acute myocardial infarction (abbreviated “AMI”) is bound to the predicate vector for CAUSES_INV_ that can be read as “is caused by.” We would expect entities of interest to have elemental vector representations similar to this bound product. The similarity between these vector representations is a natural consequence of the following steps that occur during training of the model, with diabetes mellitus (DM) as an example of an entity that meets the desired constraints:

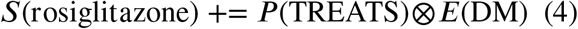

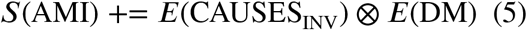

The binding operator used in the current implementation of PSI (pairwise exclusive or XOR with binary vectors as the base representation) is its own inverse, so we would expect E(DM) to be similar to both S(rosiglitazone) ⊗ P(TREATS) and S(AMI) ⊗ P(CAUSES_INV_) at the conclusion of the training process.

Recall that the semantic vector derives the meaning for each concept from the contexts (other terms and predicates) in which it has appeared. Concepts occuring in similar contexts, and therefore vector representations, will tend to have similar meanings. Concepts with similar meanings have similar vectors because semantic vectors (S) are initially zero vectors but accumulate representational information during the training. After training, the semantic vectors represent the superposition (accomplished by counting the number of set and unset bits added to the semantic vector during the training, and taking a majority vote with ties split at random) of the bound products of the elemental (E) and predicate (P) vectors representing the other concepts they occur with in semantic predications.

For the current research, the concepts that score most highly would ideally be confounders (common causes of both the drug exposure and the adverse event (outcome)). The next set of concepts in order of rank would be either upstream of the exposure or the outcome. Concepts that follow would be, to varying degrees, irrelevant.

##### Combining knowledge with data

PSI can also be used to generate discovery patterns automatically [66, 67]. Using `Semantic Vectors` syntax, we can both query the search space using discovery patterns, and generate discovery patterns from sets of paired cue terms [67, 66, 68]. In [66], PSI was used to recapitulate the discovery pattern manually constructed in [43]. While generating patterns automatically for this work, we observed that certain of these discovery patterns generated by PSI produced results suggestive of potential common causes of both the exposure and outcome - in other words, confounder candidates.

We noticed that a recurring discovery pattern is given a drug and an outcome was TREATS + COEXISTS_WITH, which suggests that rather than causing an outcome, a drug may treat a related comorbidity. (Furthermore, were it not for the related comorbidity, the patient would likely not have received the treatment, justifying the causal interpretation of the confounder “causing” the exposure).

Covariates (confounder candidates) that lie along the path of an inferred discovery pattern can be retrieved by constructing queries using the same `Semantic Vectors` syntax, as illustrated by the example confounders identified in Table 1 from querying a PSI space for concepts that relate to the (drug) allopurinol and adverse event (AE) acute liver failure in particular ways. Note that the first two confounder discovery patterns (which have only one predicate) in Table 1 only retrieve concepts that are causally related to the outcome.

**Table 1.**
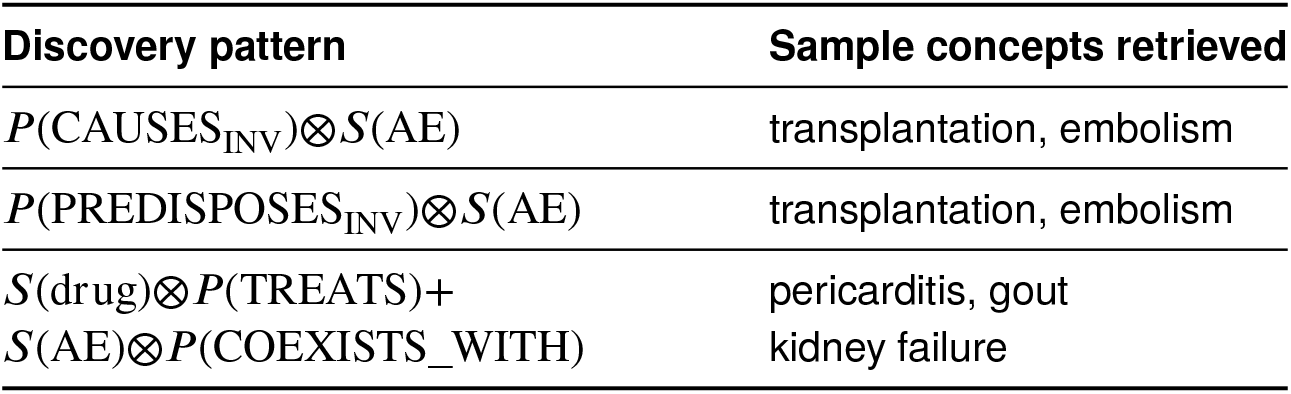
This table illustrates concepts retrieved using discovery pattern search for the drug allopurinol and the adverse event acute liver failure. “AE” = adverse event. “_*INV*_” = the inverse relation, i.e., “caused by.”

In previous work [26], we tested the extent to which literature-derived confounders could be used to accurately distinguish known causally related drug/adverse event pairs from other drug/event pairs with no known causal relationship by adjusting statistical models (multiple variable logistic regression) of data embedded in free-text clinical narrative [26]. Using the discovery patterns enumerated in Table 1, our goal was to see if including literature-derived covariates suggestive of confounding could reduce bias in data derived from free-text clinical narratives extracted from a large (de-identified) corpus of EHR data. For methodological evaluation, we used a publicly available reference dataset [69], containing labeled drug/adverse event pairs, including negative control pairs for which no relationship is known to exist. Next, we integrated up to ten literature-derived covariates into statistical models of EHR data using multiple logistic regression [26]. We used the top ten ranked confounders from PSI as a heuristic, because the top ten results generally made sense to subject-matter experts upon inspection. However, quality degraded in many cases after that rank.

We defined performance as our methodological variants’ ability to discriminate genuine causal associations from non-causal associations as labeled in the reference dataset. We measured performance by building literature-informed models instantiated with EHR data and comparing the performance of models informed by literature-derived confounders with that of naive estimates of association (e.g., reporting odds ratio and *χ*^2^). To summarize performance quantitatively, we calculated Area under the ROC (AUROC) from the ranked order of exposure coefficients from the literature-informed logistic regression models, and compared these with their *χ*^2^ baselines. Including literature-identified covariates resulted in a modest overall performance improvement of +.03 AUROC, depending on the statistical power of the available evidence used as input. A key finding was that the dual predicate discovery pattern TREATS+COEXISTS_WITH provided the most substantial performance improvement compared with single predicate discovery patterns. We reasoned that the dual predicate discovery pattern was better able to reduce confounding in the aggregate because it captures more information about both the exposure and outcome mechanisms, whereas single predicate discovery patterns only captured information about outcome mechanisms. This finding guides the choice of discovery patterns we have used in subsequent work, including the present paper.

In a follow-up study employing the same reference dataset and EHR-derived data, we again used PSI to identify causally relevant confounders to populate graphical causal models and inform causal discovery using causal structure learning methods [70]. Graphical causal models represent variables as nodes and causal relationships as directed edges. We hypothesized that the structure learning algorithm would predict fewer causal edges in light of the literature-derived confounders for the negative controls than for the reference dataset’s positive control relationships. We used the causal semantics of the predicates to orient the directed edges in causal graphs. We used the TREATS+CAUSES_INV_ discovery pattern to identify indications that are treated by the exposures and that were also noted to cause the adverse drug reactions. We picked TREATS+CAUSES_INV_ since drugs that are prescribed for an indication would likely not be taken were it not for the indication for which they were prescribed. The top-ranked literature-derived confounder candidates were then incorporated into graphical causal models. To learn graph structure, we employed the Fast Greedy Equivalence Search algorithm (FGeS) [71] implemented in the TETRAD causal discovery system [72, 73] with default algorithm hyper-parameters. FGeS is a causal structure learning algorithm that works by stochastically adding and subtracting edges until the graph’s fit for the observed data is optimized. Each drug/adverse event pair was given a score determined by the ratio of causal edges between the exposure and the outcome in the presence of all possible unique perturbations of five literature-derived confounders. Improvements in the order of +0.08 AUROC over baselines were noted from this experiment.

### 2.2. The aim of the present study

The present study documents our framework’s current stage of evolution for using computable knowledge extracted from the literature to facilitate more reliable causal inference from observational clinical data by reducing confounding bias. In our previous work, what was not clear is the extent to which the representation scheme affected the quality of the confounders and subsequently the performance of models. To probe this and other questions, we tested the following hypotheses:

- **[H1]**: that (overall) literature-informed models will reduce confounding bias in models of EHR-derived observational data and thereby improve causal inference from these data; *[premise: incorporating confounders into models should reduce confounding bias compared to naive measures of association]*
- **[H2]**: that incorporating more literature-derived confounders will improve performance over models with fewer such confounders; *[premise: models with more confounders should perform better since models with fewer confounders may result in omitted variable bias]*; and
- **[H3]**: that semantic vector-based discovery pattern confounder search (which compactly encode a vast array of information) will improve upon string-based search *[premise: a compact representation incorporating global knowledge of causal mechanisms should better prioritize information]*.

This paper builds upon an active research program for performing inference across large volumes of knowledge, though our goal is slightly different in that we are leveraging inference to infer relationships to better interpret data. This paper’s primary contributions are to highlight how background knowledge can be used to 1.) elucidate specific confounding factors and 2.) facilitate causal inference from observational clinical data in a practical setting - that of drug safety from real-world data.

## 3. Materials and Methods

### 3.1. Extracting and representing clinical narrative

We used data available from previous projects to benchmark the performance of the new approaches. Following IRB approval and a data usage agreement, we obtained permission to use the same data as in our prior studies [26, 70] from the University of Texas Health Science Center clinical data warehouse [74, 75]. These data included a large random sample of 2.2 million free-text clinical narratives recorded during outpatient encounters involving approximately 364,000 individual patients during the years 2004 and 2012 in the Houston metropolitan area.

To reveal data in the clinical narratives for downstream analysis, unstructured information embedded in the free-text narrative part of the EHR was converted to a structured format. Accurate named entity recognition (NER) is necessary for causal inference because inference requires data representations faithful to the source information. Clinical texts are known to have unique characteristics (e.g., frequent acronyms and abbreviations) that require extensive knowledge to disambiguate. Special tools are needed to process such information to take advantage of the particular characteristics of clinical language. Such issues motivate the development of clinical NER systems such as the well-regarded Medical Language Extraction and Encoding (or MedLEE, for short) clinical natural language processing (NLP) system. We preprocessed the free-text narratives using MedLEE. The original decision to use MedLEE was motivated by its performance characteristics described in the literature (see for example [76]), and the fact that internal evaluations conducted with a locally constructed reference set established that MedLEE had the best “out-the-box” performance across a range of clinical NLP tools that were publicly available at the time.

At the time the NLP extraction from the EHR was performed, MedLEE was the state-of-the-art tool for this task [77, 76, 78]. MedLEE can identify clinical concepts accurately from clinical notes, with a recall of 0.77 and a precision of 0.89 [77]. In another more recent study from 2017, MedLEE was used to detect early signs and symptoms of multiple sclerosis from clinical notes with an AU-ROC of 0.90 [0.87-0.93], sensitivity of 0.75 [0.66-0.82], and specificity of 0.91 [0.87-0.93] [79].

MedLEE encodes each concept it recognizes with a concept unique identifier (CUI) in the UMLS in a structured machine readable output format [80, 51].

To complete the unstructured data’s transformation into a structured (rectangular) dataset, we needed to extract document level concept co-occurrence statistics from the MedLEE output. To achieve this, we used the Apache Lucene [81] text indexing engine to create an index containing the normalized concepts mined from the MedLEE-processed clinical notes. We then queried the index for each concept identified in the MedLEE output, extracted document-by-concept binary arrays for each concept, and stored the resulting binary arrays in compressed files stored locally on disk. These binary arrays represent whether a concept was mentioned in a particular document. We compiled rectangular matrices by appending the resulting binary arrays (representing the presence [1] or absence [0] of a concept mentions) as our primary source of empirical data with which to test our inference procedures and instantiate our models.

### 3.2. Reference dataset

We used the popular reference dataset compiled by the Observational Medical Outcomes Partnership (OMOP) for performing a methodological evaluation of novel drug safety methods [69]. The OMOP reference dataset includes 399 drug/adverse event pairs for four clinically important adverse events.

Since the OMOP reference dataset was originally published (in 2014), varying degrees of accumulating evidence have cast doubt on specific drug/adverse event pairs’ negative control status. Hauben et al. published a list of mislabeled false negatives in the reference dataset [82] of negative control drug/adverse event pairs that have been implicated in adverse events from case reports, the literature, and pre-clinical studies. Correcting for the mislabeled false negatives noted by Hauben reduced the number of pairs for comparative evaluation. The number of pairs was reduced still further by factoring in limitations of the available empirical evidence.

Not all of the drug/adverse event pairs were well represented in the available data. We established inclusion criteria for which drug/adverse event pairs and covariates to include in our analysis by considering the implications of Peduzzi et al. [83] for the statistical power of the available EHR data. These authors studied the relationship between “events per variable” on type I and type II errors and the accuracy of variance [83]. The authors found that variables with fewer than ten events per variable are more likely to be biased. As per Peduzzi’s study, we constrained which biomedical entities or concepts (drug exposures, adverse events, or confounders) to those that occurred at least ten times. We have reported the number of drug/adverse event pairs that were compared in Table 2 (in parentheses), along with the number of drug/adverse event pairs in the original reference dataset (not in parentheses).

**Table 2.** This table presents the drug counts for each adverse event in the OMOP reference dataset. The number of drug/adverse event pairs was reduced by excluding misclassified pairs published by Hauben [82] and by the limited power of the available data in the EHR itself. The number in parentheses reflects the actual count of drug/adverse event pairs that we analyzed.

| Adverse Event Type | + Case | – Ctrl | Total |
| --- | --- | --- | --- |
| Acute kidney failure | (10) 24 | (9) 64 | (19) 88 |
| Acute liver failure | (43) 81 | (11) 37 | (54) 118 |
| Acute myocardial infarction | (15) 36 | (23) 66 | (38) 102 |
| Gastrointestinal hemorrhage | (20) 24 | (32) 67 | (52) 91 |
| Total | (88) 164 | (75) 235 | (163) 399 |

To consolidate evidence that otherwise may have been diluted across synonyms, we used the UMLS meta-thesaurus to map between synonyms of the adverse events and applied RxNorm mapping for synonym expansion at the clinical drug ingredient level. RxNorm provides a mapping from normalized names of drugs to codes used in the vocabularies of commonly used in standard clinical, pharmacological, and biomedical database applications [84]. For example, Ibuprofen’s generic concept is encoded with a UMLS concept unique identifier (CUI) string of C0020740, while the specific concept that refers to a brand-name instance of Advil Ibuprofen Caplets is C0305170. We then applied these mappings to the EHR data by using RxNorm to map from the more specific concept to the generic identifier at the pharmaceutical ingredient level.

#### Preparing SemMedDB

We downloaded and imported the latest release (at the time, version 40) of SemMedDB into a local instance of the MySQL relational database system. This version contains 97,972,561 semantic predications extracted from 29,137,782 MEDLINE titles and abstracts.

We performed several operations upon it to tailor the information it contains for the requirements of this study. For example, if a treatment is known to cause an adverse event, physicians may avoid that treatment to eliminate the potential for undesirable outcomes. To emulate what knowledge was publicly available when the reference dataset was published (2013), we excluded predicates deriving from publications after December 31st, 2012. We also removed concepts that occur ≥ 500,000 times or were considered uninformative, e.g., patients, Rattus norvegicus.

### 3.3. Searching SemMedDB for confounders

We developed and compared two variant methods for identifying a set of confounder candidates by searching computable knowledge mined from the literature (SemMedDB). Both methods apply semantic constraint search using the TREATS + CAUSES_INV_ discovery pattern but rely on distinct knowledge representation frameworks, which we refer to henceforth as “string-based” and “semantic vector-based.”

#### 3.3.1. String-based confounder search

Our first method is implemented in structured query language (SQL) and directly queries the predications table of the SemMedDB relational database. Each query takes a drug and an adverse event (called “focal concepts”) and applies the TREATS+CAUSES_INV_ discovery pattern search. The SQL query consists of two sub-queries – the first to obtain indications of the drug TREATS and the second to obtain the indications that also cause the adverse event. The result set should contain a list of confounder candidates that fulfill both of these semantic constraints.

To find the best subset of confounders, we developed a score to rank the confounders by the strength of their support as confounders in the literature. To score confounders, we calculated the product of the counts for each confounder given the number of citations from each arm of the discovery pattern query (the TREATS arm and the CAUSES_INV_ arm). Results were next ranked in descending order of this product score. To screen out potential errors from machine reading, concepts with less than two mentions were excluded from the result set.

#### 3.3.2. Semantic vector-based confounder search

Our second method transforms SemMedDB using PSI, the distributional representation scheme introduced earlier, that facilitates vector algebra inference (for “vector-based” confounder search). Using the truncated version of SemMedDB described above as input, we derived a binary PSI space with 32,000 dimensions (in bits). We used inverse document frequency weighting to adjust for frequently occurring but uninformative predications and removed generic terms with little discriminative meaning (included in Appendix A).

### 3.4. Additional confounder candidate filtering procedures

We conservatively mapped between the reference dataset and concepts from the UMLS and between observed variables represented in the EHR data and knowledge represented by concepts in the UMLS. Confounder candidates were only incorporated into models if they appeared in the EHR data at least ten times with both the exposure and the outcome variables (following the heuristic of Peduzzi et al. [83]). Additional filters and criteria were applied for the inclusion of confounder candidates in the models. For instance, we identified highly frequent but uninformative stopwords such as “disease” and “gene” (see Appendix A). We also excluded concepts that were synonyms of the drug exposure and adverse event, as these terms arise from search procedures, sometimes either from machine reading errors or the confounder search procedure itself. The exclusion of synonymous concepts was achieved by checking to see if the confounder candidate’s preferred name in the UMLS matched with the strings representing the drug exposures and adverse event outcomes. Confounders were excluded if they matched either the exposure or the outcome.

To evaluate the effect of adding different amounts of literature-derived confounders, we applied two thresholds on the number of confounders: a lower threshold of five and an upper threshold of ten. In the next paragraphs, we have explained our reasoning for why we chose these thresholds.

String-based search can only yield results that match strings from user input. In contrast, the continuous nature of the underlying vector-based knowledge representation determines that all of the entities represented in that vector space are related to varying degrees.

To limit the number of search results, there are two primary methods: by either applying an arbitrary cut-off from the distributional statistics or applying a threshold that limits the number of search results. The first method for narrowing search results operates by setting a cut-off on the number of standard deviations required to be included in the search results given the mean similarity scores from the PSI query. In our internal evaluations, we attempted to use a threshold of 2.5 standard deviations to constrain the number of confounders. However, we found that this approach had detrimental effects, including 1.) uneven quality of results (standard deviation was not sufficient in some cases to screen out irrelevant noise); 2.) inconsistent numbers of results at that threshold (sometimes many quality confounders, other times, PSI would filter out seemingly relevant confounders). That is, working with statistical thresholds proved difficult as the utility of this threshold varied depending on the query. Accordingly, we chose not to use a standard deviation based threshold, and reverted to the second method for limiting results.

For our research problem, we were left with a heuristic strategy for how to compare the effect of not only different search and representation strategies (string-based vs. semantic vector-based confounder search) and modeling methods but testing the effects of incorporating different amounts of information, i.e., numbers of literature-derived confounders.

In our previous experience using the literature to identify confounder candidates, we observed that roughly the first ten concepts returned from our confounder search were medically meaningful. Hence, we picked ten for the upper threshold. To simulate omitted variable bias, we divided ten in half to arrive at the lower threshold of five confounders. Thus, there is a reasonable expectation that roughly comparable numbers of valid confounder candidates can be identified within these thresholds from either string-based and semantic vector-based confounder search can be integrated into the models.

### 3.5. Assembling models from knowledge and data

As input, concept-by-observation matrices were constructed for each drug/adverse event in the reference dataset for which we possessed sufficient data. The input matrices for each drug/adverse event pair consist of columns representing the drug exposure, adverse event outcome, and confounder concepts (where the confounder concepts have been identified using one of the confounder search methods described above), while each row represents the presence or absence of concept mentions extracted from the narratives in clinical notes.

#### 3.5.1. Literature-informed regression modeling

We applied off-the-shelf multiple variable logistic regression to the EHR data, adjusting for the literature-derived confounders, where Y = outcome (the adverse event), A = exposure (drug), W = the set of confounders, with the Greek letters {*α*}, {*β*}, and {*γ*} representing the intercept, the regression coefficients of the exposure and the covariates, respectively:

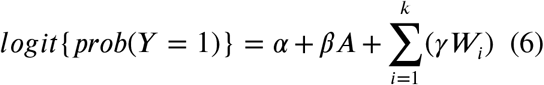

In this paper, we use logistic regression as a comparator method to the exact causal inference method introduced in the next subsection. Logistic regression provides “guardrails” on more advanced methods since regression usually does a decent job at adjustment and provides a predictive check on the adjustments. For further discussion on the relationship between causal effect estimation and regression coefficients, see Chapter 6 in [85].

#### 3.5.2. Literature-informed graphical causal modeling

To construct graphical causal models, we used the **bnlearn** R package [86]. The **bnlearn** package allows the user to incorporate variables and define the relationships between variables. We exploited structural information from the literature to create “white lists” (lists of required edges) and “black lists” (lists of prohibited edges) to orient those edges. The white lists contain mandatory labeled edges between each of the confounders and the drug and adverse event, while black lists forbid effects from causing drug exposures.

To learn dependency relationships additional to those identified from the literature, we applied the Max-Min Hill-Climbing (MMHC) algorithm, first described by Tsamardinos et al. [87] and implemented in **bnlearn** [86] with default hyper-parameter settings. MMHC is a hybrid structure learning algorithm that first uses a constraint-based search to learn the dependency structure from data and then orients the edges of the graph using the a score-based search algorithm that finds a structure that best fits the data and background knowledge by optimizing a criterion score. We used the Bayesian Information Criterion score described in [88]. Next, we applied the maximum likelihood estimation (MLE) procedure within **bnlearn** to find the configuration of weights associated with each edge in the graph given the data and the graph structure. These weights quantify the strength of the dependencies between variables. Once the structure and the weights have been learned, the model is ready to answer questions of interest.

To estimate causal effects, we applied the classic exact inference procedure known as the junction tree algorithm, as implemented with default settings in the R package **gRain** [89]. The junction tree algorithm efficiently computes posterior probabilities by transforming the DAG into a tree structure. The resulting tree structure then propagates updated values using the sum-product method across the graph. In our use case, the end goal of updating values is to compute the potential outcomes at different levels of exposure.

The junction tree method is reasonably efficient because the graph is sparse (unsaturated), and all calculations are local. Next, we query the resulting object by telling it to “listen” to the adverse event node in the graph, and by fixing the value of the exposure/treatment to “1” and then to “0,” and then subtracting the difference to obtain the ATE (Δ) (see Equation 1). More details about the junction tree algorithm’s derivation may be found in [90, 91].

### 3.6. Overview of the evaluation framework

The steps of our modeling and evaluation framework are outlined below and illustrated in Figure 3:

**Figure 3:**
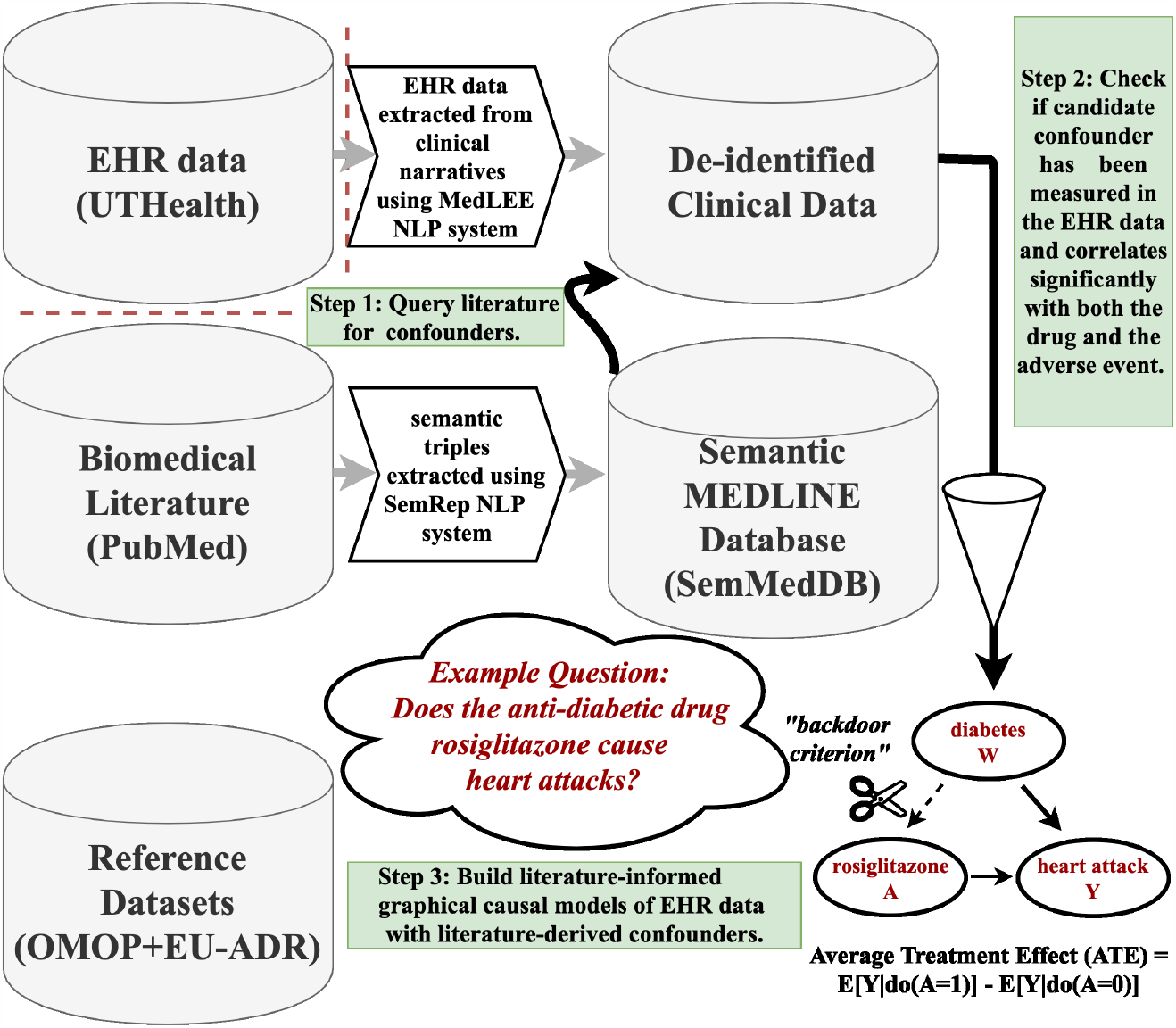
Illustration of our literature-informed modeling framework.

1. Query the literature for confounders using either string-based or semantic vector-based literature search.
2. Determine the eligibility for the inclusion of each confounder candidate in the order of its retrieval. Test each confounder for at least ten co-mentions with both the drug and the adverse event using the clinical data and stop testing after obtaining the thresholds of five or ten confounding variable candidates, or until reaching the end of the list of the confounder candidates, if less than the thresholds, when using string-based search.
3. Build statistical and graphical causal models incorporating varying numbers of literature-derived confounders to predict and estimate causal effects from the EHR-derived empirical data.

We evaluated the performance of our modeling procedures by comparing the relative performance of naive measures of association (ROR, and *χ*^2^) with statistical and causal models informed by the literature and instantiated with EHR-derived data. The EHR data encode concept mentions with discrete dichotomous (binary) variables. We used the reference dataset labels as ground truth.

Different drug/adverse event pairs can be expected to have a range of effect sizes, with the true effect sizes of interest (as one might collect under randomization) ranging across varying intervals. Two following two assumptions underlie our methodological comparison:

- **ASSUMPTION I**: the causal effect estimates and adjusted measures of association should be greater in magnitude for true positive drug/adverse event pairs than the causal effect estimates and adjusted measures of association for negative drug/adverse event pairs in the reference dataset; and
- **ASSUMPTION II**: the effect estimates and adjusted measures of association of the negative controls should approach zero.

Working under the above assumptions, we measured performance by calculating Area under the Curve of the receiver-operating characteristic (AUROCs), Area under the Precision and Recall Curve (AUPRC), and Mean Average Precision at K (MAPK) from the ranked ordering of the following statistics: **baselines**: [*χ*^2^, reporting odds ratio (ROR)] and **modeling scores**: the coefficients from performing multiple logistic regression (*β*) and average treatment effects (ATE) from the graphical causal models (Δ).

To evaluate our literature-informed confounding variable identification framework, we aggregated performance statistics across the following methodological variants:

- **more (literature-derived) information vs. less**
- **string-based vs. semantic vector-based search**

Finally, to obtain insight into overall performance, we needed to consolidate performance statistics across adverse event types, albeit at the expense of fine detail since each adverse event has a different underlying prevalence and other unique characteristics. To calculate global summary scores, we normalized the scores by transposing them onto a similar scale. We weighted the scores (whether *β* or Δ) for each adverse event type by computing the proportion of the total number of pairs contributed to the total score from each adverse event type and then multiplied the individual scores for each drug / adverse event pair by that proportion. The weighted scores were then compiled into overall weighted metrics.

### 3.7. Software infrastructure

The PostgreSQL relational database system and various R statistical packages were used to analyze this study’s data. The list below enumerates that software packages used for this study: **R base** version 3.6, **gRain** version 1.3-3: exact causal inference [89], **ggpubr** version 0.4.0.999 for boxplots, **RPostgreSQL** version 0.6-2: library for R connectivity with Postgres relational database, **pROC** version 1.16.1: ROC curves, **PRROC**: Precision-Recall curves, **tidyverse** version 1.3.0: data manipulation [92], and **bnlearn** version 4.5: graphical modeling and parameter estimation [93, 86]. We also used `Semantic Vectors` version 5.9 [63, 64] (running Oracle Java 1.8.0_231).

The software and more extensive information and models developed for this paper are publicly available on the causalSemantics GitHub repository. The Material on GitHub includes Receiver-Operator Characteristic (ROC) and Precision-Recall curves along with the data tables, box plots comparing the various procedures, sample visualizations of the causal graphical models, and confounder sets for the drug/adverse event pairs from the reference dataset.

## 4. Results

We begin by enumerating high-level statistics and then proceed to tease out the strengths and weaknesses based on what the metrics are telling us about each of the methodological variants, we break down the results from comparing our modeling procedures in terms of each performance metric. Then we will consider in the discussion the implications for our hypotheses and future work and conclude with the lessons learned.

The prevalence of each adverse event in our EHR data was as follows: acute kidney failure: 4,847; acute liver failure: 28,217; acute myocardial infarction: 54,550; and gastrointestinal hemorrhage: 59,695.

We analyzed the number of confounders from string-based search per drug / adverse event pair in the reference dataset. The mean number of confounders per pair was ten, and the median was approximately seven. The minimum was 0, and the maximum was 121.

Tables 3, 4, and 5 show the results for various performance metrics providing different perspectives (AUROC, AUPRC, and MAP-K, respectively) on performance. The AUROC in Table 3 provides a global assessment of classifier performance irrespective of the classification threshold, while AUPRC in Table 4 is preferred with imbalanced reference datasets [94]. Table 5 presents MAP-K, which considers the top-ranked results, and is arguably the most important metric for practical purposes.

**Table 3.** This table shows AUROC scores from the various models. *χ*^2^ = chi squared. *β*= coefficient from multiple variable logistic regression. Δ= average treatment effect using graphical causal models. Results exceeding best baseline performance are in *boldface*. † indicates best performance for a side effect.

|  | Baselines |  | Statistical Models |  |  |  | Causal Models |  |  |  |
| --- | --- | --- | --- | --- | --- | --- | --- | --- | --- | --- |
| Adverse event type [+ ctrls, - ctrls] | ROR | $\chi^2$ | $\beta_{sql}^5$ | $\beta_{psi}^5$ | $\beta_{sql}^{10}$ | $\beta_{psi}^{10}$ | $\Delta_{sql}^5$ | $\Delta_{psi}^5$ | $\Delta_{sql}^{10}$ | $\Delta_{psi}^{10}$ |
| Acute kidney failure [10+, 9-] | 0.6222 | 0.4667 | 0.6 | **0.7444** | 0.5667 | 0.6222 | 0.5 | **0.8111†** | 0.5111 | 0.5 |
| Acute liver failure [43+, 11-] | 0.5835 | 0.5814 | **0.6004** | **0.6723** | 0.4905 | **0.6765†** | 0.4926 | 0.5243 | 0.5433 | 0.5624 |
| Acute myocardial infarction [15+, 23-] | 0.6261 | 0.6754 | 0.658 | 0.5159 | 0.4783 | 0.5217 | 0.6 | 0.6725 | 0.6667 | 0.6464 |
| Gastrointestinal hemorrhage [20+, 32-] | 0.5688 | 0.6 | 0.5672 | **0.7078** | 0.5688 | **0.7156†** | 0.5484 | 0.55 | 0.5203 | 0.4891 |
| Weighted overall AUROC [88+, 75-] | 0.5933 | 0.5959 | **0.6032** | **0.6556†** | 0.5363 | **0.6005** | 0.5215 | **0.6466** | 0.561 | 0.5513 |

**Table 4.** Area under the Precision Recall Curve (AUPRC).

|  | Baselines |  | Statistical Models |  |  |  | Causal Models |  |  |  |
| --- | --- | --- | --- | --- | --- | --- | --- | --- | --- | --- |
| Adverse Event Type [+ ctrls, - ctrls] | ROR | $\chi^2$ | $\beta_{sql}^5$ | $\beta_{psi}^5$ | $\beta_{sql}^{10}$ | $\beta_{psi}^{10}$ | $\Delta_{sql}^5$ | $\Delta_{psi}^5$ | $\Delta_{sql}^{10}$ | $\Delta_{psi}^{10}$ |
| Acute kidney failure [10+, 9-] | 0.4415 | 0.47 | 0.4316 | 0.3832 | 0.4535 | 0.4276 | **0.4889** | **0.6785†** | **0.4781** | **0.5032** |
| Acute liver failure [43+, 11-] | 0.7554 | 0.7562 | 0.7464 | 0.7057 | **0.7726** | **0.8055** | **0.7875** | 0.7028 | 0.7303 | **0.8281†** |
| Acute myocardial infarction [15+, 23-] | 0.3387 | 0.5277 | 0.4816 | 0.3743 | 0.4018 | 0.3735 | 0.4499 | **0.5429** | **0.5631†** | 0.4943 |
| Gastrointestinal hemorrhage [20+, 32-] | 0.3243 | 0.308 | 0.3192 | 0.2744 | 0.3189 | 0.2735 | **0.3694** | **0.3773†** | **0.3344** | **0.3397** |
| Weighted overall AUPRC [88+, 75-] | 0.4841 | 0.5266 | 0.5117 | 0.4533 | 0.5092 | 0.457 | **0.5357** | **0.5929†** | **0.5356** | **0.5566** |

**Table 5.** Mean Average Precision at K. ROR = reporting odds ratio. *χ*^2^ = chi squared. *β*= generalized linear models (multiple variable logistic regression). Δ= average treatment effect using graphical causal models. Results exceeding best baseline performance are in *boldface*. † indicates best performance for a side effect.

|  | Baselines |  | Statistical Models |  |  |  | Causal Models |  |  |  |
| --- | --- | --- | --- | --- | --- | --- | --- | --- | --- | --- |
| Adverse Event Type [+ ctrls, - ctrls] | ROR | $\chi^2$ | $\beta_{sql}^5$ | $\beta_{psi}^5$ | $\beta_{sql}^{10}$ | $\beta_{psi}^{10}$ | $\Delta_{sql}^5$ | $\Delta_{psi}^5$ | $\Delta_{sql}^{10}$ | $\Delta_{psi}^{10}$ |
| **K = 10** |  |  |  |  |  |  |  |  |  |  |
| Acute kidney failure [10+, 9-] | 0.4756 | 0.4921 | 0.4106 | 0.3068 | 0.4768 | 0.4135 | **0.5378** | **0.7032†** | 0.4756 | **0.5653** |
| Acute liver failure [43+, 11-] | 0.8955 | 0.8917 | 0.8389 | 0.7579 | 0.8803 | 0.725 | 0.7972 | 0.8951 | 0.6815 | **0.9627†** |
| Acute myocardial infarction [15+, 23-] | 0.4889 | 0.5981 | 0.4974 | 0.4021 | 0.4911 | 0.4155 | 0.5193 | **0.8529†** | **0.7296** | 0.5883 |
| Gastrointestinal hemorrhage [20+, 32-] | 0.5556 | NA | NA | NA | 0.1 | 0.125 | NA | 0.325 | 0.125 | 0.2 |
| **K = 25** |  |  |  |  |  |  |  |  |  |  |
| Acute kidney failure [10+, 9-] | 0.4866 | 0.515 | 0.4751 | 0.4166 | 0.5057 | 0.4665 | **0.5422** | **0.7277†** | **0.5229** | **0.5477** |
| Acute liver failure [43+, 11-] | 0.7993 | 0.7863 | 0.7767 | 0.7132 | **0.8449** | 0.7067 | **0.8163** | **0.8764** | 0.7085 | **0.9051†** |
| Acute myocardial infarction [15+, 23-] | 0.3795 | 0.6056 | 0.54 | 0.4227 | 0.455 | 0.4261 | 0.5092 | **0.6715†** | **0.6595** | 0.5618 |
| Gastrointestinal hemorrhage [20+, 32-] | 0.4258† | 0.2093 | 0.224 | 0.1937 | 0.255 | 0.1857 | 0.3025 | 0.3739 | 0.2361 | 0.2626 |

Although the subset of the reference dataset we analyzed was only moderately imbalanced overall, the class imbalance for each particular adverse event ranged from approximately balanced (acute kidney failure - with ten positive cases and eleven negative control drug/adverse event pairs) to strongly imbalanced (acute liver failure - with forty-three positive cases and eleven negative control drug/adverse event pairs). Note that because of class imbalance where there are more positive event pairs than negative, we refrain from giving equal weight to the performance of acute liver failure, though we report it in the results.

### AUROC

As shown in Table 3, the spectrum of performance was broad. The methodological variant that performed well the most consistently was 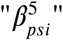 (or the variant using up to five semantic vector-based confounders with multiple logistic regression), followed by 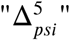 (or the variant using up to 5 semantic-vector-based confounders calculating the ATE), as shown in Figure 4 illustrating unweighted overall AUROCs. Improvement over the baseline was most consistent for gastrointestinal hemorrhage. Vector-based models bested string-based models. More vs. fewer confounders was a draw. Logistic regression estimation performed better than ATE. We included the scores from acute liver failure in Figure 4 because the overall class imbalance was extreme; there were 88 positive cases and 75 negative controls, as per Table 2.

**Figure 4:**
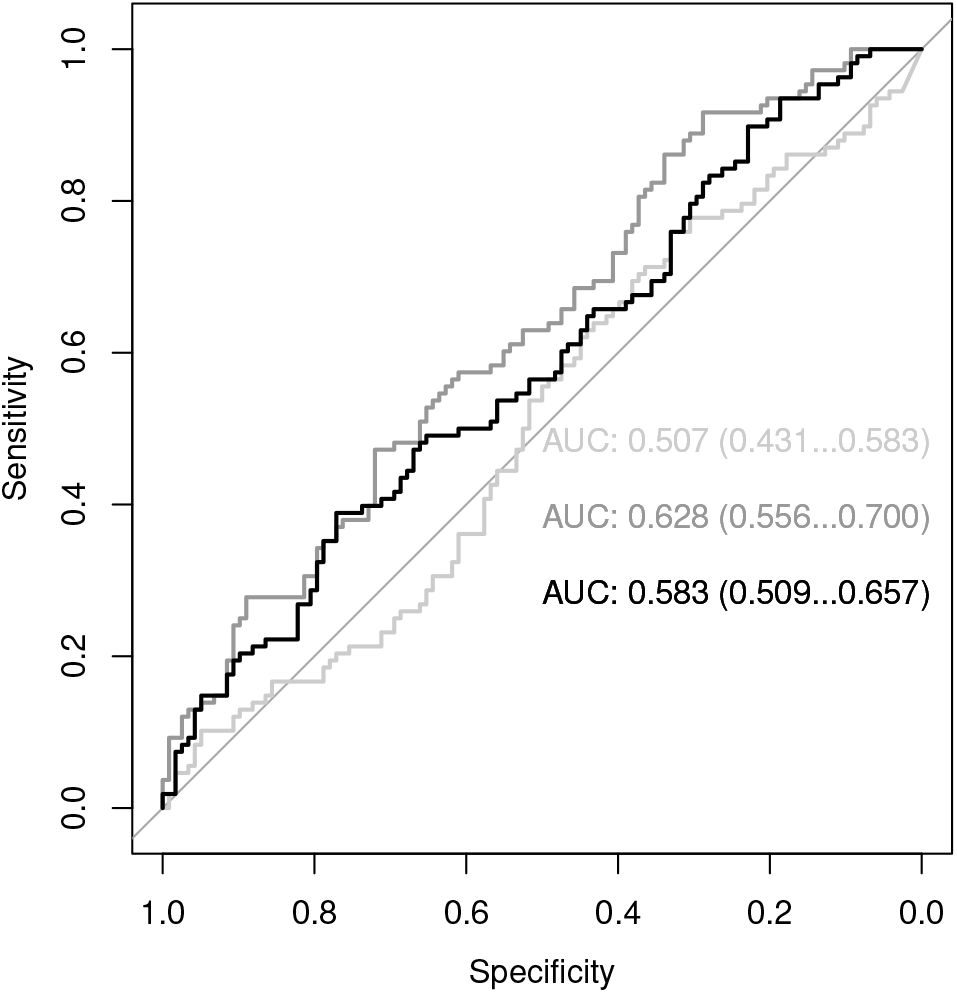
Unweighted AUROC for best results for literature-derived confounders (querying method = PSI, # of confounders = 10) vs. (baseline = *χ*^2^). Light gray = *χ*^2^. Medium gray = [logistic regression coefficient]. Black = [average treatment effect/Δ]. The numbers to the right of the ROCs represent the 95% confidence intervals.

### AUPRC

AUPRC is preferred in analyzing classification problems where the classes are not well balanced between positive and negative labels. AUPRC scores range from 0.0 being the worst to 1.0 being the best, unlike AUROC scores, which usually range from 0.5 to 1.0. Acute kidney failure had the most improvement in the AUPRC metric with confounding adjustment.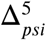 performed well the most consistently. Vector-based models bested string-based models. More vs. fewer confounders was a draw. ATE performed better than Logistic regression.

### MAP-K

As one might anticipate with methods intended to correct for false positives induced by an otherwise unmeasured confounding effect, consistent performance improvements with adjustment are found with the MAP-K metric, which measures the accuracy of the top-ranked (most strongly predicted) results. Arguably, MAP-K is the most important metric for prioritizing signals, the primary application focus of this paper. The importance of MAP-K is apparent when comparing overall performance between the best baseline and best-adjusted models, with improvements of 0.05 and 0.1 with k=10 and 25, respectively. The best performance was observed with the 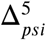 models, which scored the highest MAP-K with k=25. Vector-based models bested string-based models. More vs. fewer confounders was a draw. ATE performed better than Logistic regression.

### Performance Summary

While performance differs across metrics, adjustment using the literature-derived confounders improved predictive and causal inference performance over naive baselines of association across all four adverse events. There were substantive improvements for particular adverse events, with increases in AUROC of 0.1 and 0.2 over the best baseline model with the best-adjusted models for gastrointestinal hemorrhage and acute kidney failure, respectively, and smaller but consistent improvements in AUPRC where best performance was always attained by one of the adjusted models. considering the class imbalances of each condition and the importance of MAP-K, The best performance was most frequently attained by 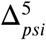 models. In general, semantic vectors-based models bested string-based models, fewer confounders bested more (though not always, particularly using the MAP-K metric), and Δ bested *β* estimates.

## 5. Discussion

### [H1]: Does literature-informed modeling reduce bias?

In most cases, the adjusted models show performance improvements over the unadjusted baseline measures of association. While there was a substantial reduction of bias, there was room for improvement. The overall improvement was consistent with but not significantly better than that from previous work [26, 70].

We analyzed the distribution of the Δs and *β*s across the methodological variants. Across all adverse events, the mean Δs for the positive controls were higher than for the negative controls in the reference dataset. For example, for 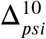, the mean Δ for the negative controls was 0.03 (for reference, the mean Δ of the positive controls was 0.05). An ideal deconfounding method would reduce the Δs for the negative controls to zero. This was not the case with certain cases, where the mean adjusted scores for the positive cases were lower than the negative controls, thus violating ASSUMPTION II mentioned earlier. We explore this issue in more detail below.

### [H2]: Does adding more literature-derived confounders versus fewer improve performance?

For two out of three metrics, as per Tables 4 and 5 (remembering that acute liver failure is problematic in its level of class imbalance towards the positive), many of the high-performing methodological variants were those with five literature-derived confounders, though that was not always the case. From the standpoint of causal theory, what may be happening is that the additional confounder candidates at the higher threshold are introducing overcontrol bias [95] and in other cases the additional confounders address omitted variable bias, resulting in better performance [96]. Thus, the extent to which adding more literature-derived confounders is inconclusive, and likely depends on the particular local causal structure and quality of data in each case.

### [H3]: Which confounder search method (string-based versus semantic vector-based) results in better performing models?

We can observe interesting patterns comparing the individual adverse drug reaction summary results of string-based or semantic vector-based confounder search.

With few exceptions, models informed with confounders from semantic vector-based confounder search performed better than models informed with confounders from string-based confounder search. We expected the string-based search’s conservative nature to result in missing coverage of many drug/adverse event pairs. In many cases, this suspicion was confirmed when no confounders were available after screening out synonyms of the exposure and outcome and stopword-like concepts for the biomedical domain, e.g., patients, therapeutic procedure, Rattus norvegicus).

It is clear that how knowledge is represented and organized can affect measures of topical relevancy, which in turn affect the specific set of confounder candidates retrieved by a query. The question of how the representation of knowledge affects model performance is an important one. It also follows that the quality of the confounders affects the ability of the method to reduce confounding.

The present paper partially lends supportive evidence for what Vanderweele calls the disjunctive cause criterion, or DCC [97, 5]. The DCC is a criterion for selecting covariates for which to adjust and recommends selecting known determinants of either the exposure or the outcome, or both. Arguably, a major factor bolstering the performance of semantic vector-based search is the proportion of cue concept (drug or exposure) contexts occupied by the target (confounder) concept in the underlying knowledge representation.

To the extent that PSI can pick either determinants of the exposure or the outcome or both, our framework is a step toward implementing the DCC for (partially)-automated causal inference. In contrast to the relatively brittle Boolean confounder search used with string matching, the Hamming distance metric used to measure the similarity between vector space representations is continuous in nature, permitting partial match when only one of the two constraints is met.

To extrapolate from our results, a key factor bolstering the performance of semantic vector-based search lies in its knowledge representation. The advantage of using semantic vector-based search over string-based search is analogous to the advantages offered by Salton’s original vector space model as an alternative to Boolean retrieval when it was first introduced [98, 99].

Recall that the median count of confounders per drug/adverse event pair in the reference dataset (from string-based search) was approximately seven. This means that string-based search was limited by whether or not it could find an exact match in SemMedDB.

The finding of improved performance is perhaps not surprising, given the development effort that has improved PSI’s ability to perform approximate reasoning over large bodies of knowledge. One explanation for why semantic vector-based confounder search performed better than string-based search is that semantic vector-based search can better integrate relevant information more flexibly: because semantic vector-based search quantifies results on a continuous scale, unlike the exact matching underlying the basis of string-based search. Another explanation for why semantic vector-based search usually performs better than string-based search is that semantic vectors are normalized. Normalization prevents frequently occurring concepts from dominating the result set.

All aside, a possible research direction would be to combine the results of both string-based and semantic vectors-based confounder candidates for adjustment or simply to use explicit co-occurrence as a constraint on semantic vector search.

In brief, string-based search takes a verbatim interpretation of the request and provides results as specified by the query (“what the user says they want”), whereas semantic vector-based search applies vector algebra to infer in a sense what the user truly *needs*.

#### Comparing estimation methods

Literature-derived computable knowledge was found to be useful for informing causal inference (as implicated by the performance metrics). (Δs) from the graphical causal models bested Logistic regression models (*β*s).

To connect prediction and causal inference (estimation), a less biased estimate of the drug effect is likely to lead to better prediction performance. While prediction and causal inference tasks are closely related, they are not the same: prediction optimizes by minimizing variance, whereas the objective of causal inference problems is to reduce or eliminate bias. Nevertheless, using regression with known confounders as regressors is a traditional way of performing causal inference, where the *β* in Equation 6 has been interpreted as the causal effect [100, 101, 102].

#### Causal graph example

We have included a sample graph in Figure 5. This figure provides a sample of the structure and content expressively modeled by the literature-informed graphical causal graph formalism and instantiated with EHR-derived observational clinical data from free-text clinical narratives. Noting the centrality of asthma in the graph, we searched the literature to find that asthma as an indication is associated with a two-fold increased risk of AMI. While inactive asthma did not increase the risk of AMI, individuals with active asthma had a higher odds of AMI than those without asthma (adjusted OR: 3.18; 95% CI: 1.57 - 6.44) [103]. More such graphs are available in the GitHub repository.

**Figure 5:**
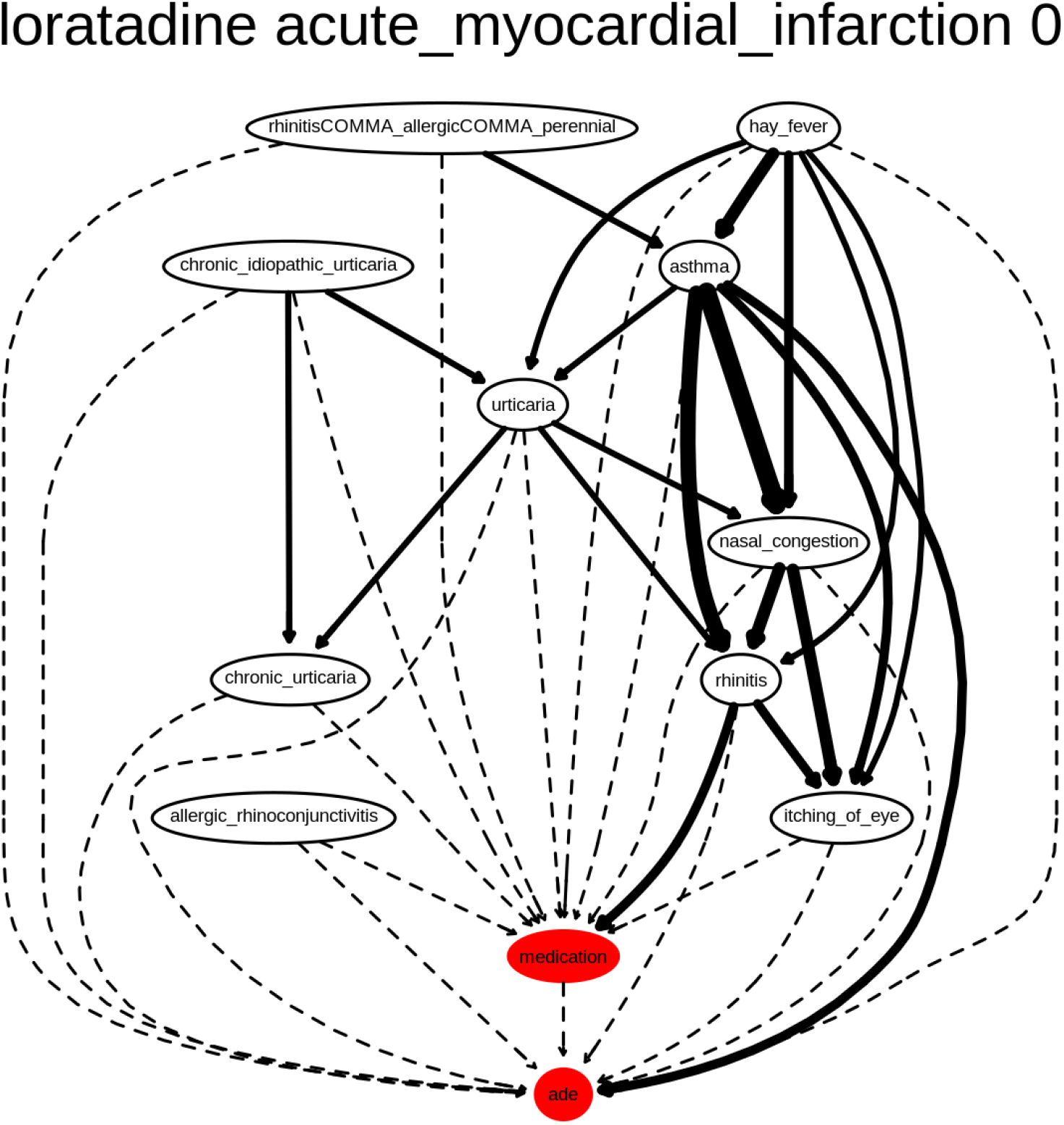
Graphical causal model using PSI at the 10 confounder threshold for loratadine, a negative case for acute myocardial infarction. The thickness of the edges indicates the strength of the observed relationship in the EHR-derived data.

#### 5.1. Comparison with previous related work

For the sake of a coarse comparison, the AUROCs of several EHR-based pharmacovigilance methods have been included in Table 6. However, note that the performance patterns are not strictly comparable owing to different sample sizes and populations. However, these results have been included here for convenience and to provide context, however crude.

**Table 6.** This table shows AUROC statistics from several EHR-based drug safety studies. Because these studies used different subsets of the reference dataset, the comparisons are not strictly comparable. Also note that all of the studies used exclusively EHR data as the primary source of empirical data, except for [104], which incorporated other sources of empirical data including claims and FAERS-derived data to attain a higher adjusted score, pointing to a promising potential area of future drug safety research.

| Study | Li (2014) | Li (2015) | Malec (2016) | Malec (2018) | This study |
| --- | --- | --- | --- | --- | --- |
| Naive | 0.53 | 0.51 | 0.53 | 0.504 | 0.55 |
| Adjusted | 0.51 (lasso) | 0.89 (meta-analysis) | 0.55 | 0.58 | 0.65 |

In summary, our literature-informed graphical causal modeling framework resulted in superior performance compared with our previous purely EHR-based modeling efforts [26, 70] but fare poorly in comparison with results from applying meta-analysis from [104] with an AUROC of 0.89, as seen in the table 6. However modest the improvement, the success of such a principled approach using causal models applied to coarse cross-sectional data opens many doors for methodological refinement and future avenues of research.

For example, incorporating outside statistical information (e.g., from FAERS and claims data) resulted in more considerable performance improvements than could be obtained with confounding correction alone. Our approach could be readily applied to any of these data sources or their combination also.

#### 5.2. Practical applications of literature-informed modeling

Domain knowledge improves the efficiency of causal learning tasks by:

- **Reducing the dimensionality of features**: the richness of EHR data introduces the “curse of dimensionality” problem, presenting a large number of potential covariates for which to adjust. By contrast, discovery pattern search can provide a parsimonious set of covariates vetted from background knowledge that is useful in many situations for explaining, controlling for, and reducing confounding bias. That is, EHR data have a large number of variables and are, hence, high dimensional; having computable domain knowledge permits the efficient identification of a feasibly-sized set of confounders.
- **Simplifying causal structure**: qualitative information about the orientation of variables in causal graphs simplifies the task of learning causal structure.
- **Providing *a priori* knowledge of causal order**: although time is not coded explicitly in cross-sectional data, *a priori* knowledge provides information about the likely ordering of events. In terms of graphs, when we assume that a biomedical entity is a confounder relative to exposure and an outcome, then it has a set topological structure (probabilistic and causal dependency). Without assumptions drawing from substantive knowledge, it is often impossible to determine the causal direction (such that it exists) from the data alone [34], though much progress is being made in this area [105, 6].

Discovery pattern search could be useful for identifying variables with causal roles besides that of being a confounder. For example, investigators may also be interested in identifying certain types of variable, such as:

- **Colliders**, or common effects of both the exposure and outcomes variables, that can amplify bias [106, 3, 107]; and
- **Mediators**, or intermediate variables which lie along the causal chain from the exposure to the outcome (and which can also bias estimates if a variant of “total effect” is the causal parameter of interest) [108].

While we have not screened for such variables in the current paper, we present the discovery patterns in Table 7 as a starting point for future research.

**Table 7.** Discovery patterns for identifying different variable types.

| Variable role | Query | Graph shape |
| --- | --- | --- |
| **outcome mechanisms** | $X \text{CAUSES}_{\text{INV}} AE$ | $AE \leftarrow \text{outcome\_mechanisms}$ |
| **Mediators** | $\text{drugCAUSES} X;$<br>$X \text{CAUSES}_{\text{INV}} AE;$ | $\text{drug} \rightarrow \text{mediator} \rightarrow AE$<br>$\text{drug} \rightarrow \text{mediator} \rightarrow AE$ |
| **Colliders** | $\text{drugCAUSES} X;$<br>$AE \text{CAUSES} X;$ | $\text{drug} \rightarrow \text{collider} \leftarrow AE$<br>$\text{drug} \rightarrow \text{collider} \leftarrow AE$ |

Furthermore, although we have only used the TREATS and CAUSES-derived predicates in this study, other potentially useful “causal” predicates, and related discovery patterns, exist. For example, other likely useful predicates include: PREDISPOSES, AFFECTS, STIMULATES, PREVENTS, INHIBITS, and PRODUCES.

Note that the discovery patterns we have used in our paper are “manually-designed” discovery patterns. As mentioned earlier, methods exist that can automatically generate discovery patterns directly from the distributional semantics of PSI spaces [66, 67] but exploring other discovery patterns is beyond the scope of the present paper.

#### 5.3. Error analysis

We interrogated our modeling procedures to try to identify why they failed and to gather ideas on how to address these issues in subsequent work. We bring the case of ketorolac and acute myocardial infarction to attention. Initially, we thought that the listing of the right platysma (a facial muscle) demonstrated PSI’s power to infer relationships based on the global similarity of structured knowledge. Although there were no results in PubMed or SemMedDB linking “right platysma” to ketorolac and acute myocardial infarctions, Keterolac has been a useful adjunct to Botox treatment to reduce discomfort (after facial injection). Ketorolac was also studied in an RCT for biliary colic pain [109].

However, we note that PSI can only draw inferences based on the global similarity of structured knowledge when deliberately asked to (using e.g.,two-predicate path queries from semantic vector cue to semantic vector target). The results of the current search method do not benefit from vector similarity, as we are retrieving elemental vectors, which are, by definition, dissimilar. Another explanation could be that this suggestion may have resulted from the random overlap between elemental vectors. Within our broader methodological framework, it is helpful that the empirical data can often be used to correct for machine reading or information retrieval errors in causal models. By contrast, string-based search often yielded confounder candidates that were overly general. Confounder search yielding generic confounder candidates was less often the case with PSI, presumably because statistical weighting was used to deliberately limit the influence of frequently occurring terms during the construction of the vector space.

As per table 8, the explanation we considered for the poor performance we observed in some cases was that some of the TREATS relationships mined by SemRep in SemMedDB were occasionally more suggestive of potential non-standard uses that are not as yet FDA approved. It would be unlikely that a patient would be prescribed the drug for that particular (confounder candidate) indication, though it could still affect the outcome. Also, we noted other cases of hedging, the use of case reports, and anecdotal evidence.

**Table 8.** Problematic source sentences. AKI = acute kidney failure.

| Problem | Source → Target | Confounder | Source sentence |
| --- | --- | --- | --- |
| Speculation | ibuprofen →<sub>TREATS</sub> X | Benign prostatic hypertrophy | (PMID: 15947693) 'Further research must be done to investigate the potential use of ibuprofen in patients with BPH and examine if JM-27 expression in patients with BPH may stratify individuals who may be most responsive to pharmacological treatment.' |
| Negative evidence | ketoconazole →<sub>TREATS</sub> X | Hypercalcemia | (PMID: 11033844) 'However, deterioration of renal function during ketoconazole administration as well as failure of hypercalcemia to be affected during short-term ketoconazole treatment suggest that this drug might not be appropriate for acute treatment of hypercalcemic sarcoidosis.' |
| Hedging | X →<sub>CAUSES</sub> AKI | Toxic Epidermal Necrolysis / Steven's-Johnson Syndrome | (PMID: 19155617) 'CONCLUSION: AKI, the need for dialysis, and late hypokalemia could be the consequences of SJS/TEN.' |
| Hedging | ketoconazole →<sub>TREATS</sub> X | Tuberculosis | (PMID: 17644711) 'Further investigation is necessary to determine the role of KTC in the treatment of TB.,' '17644711' |
| Case report | X →<sub>CAUSES</sub> AKI | Tuberculosis | (PMID: 2386602) 'A 78 year old male patient who had been treated by haemodialysis for 17 years for renal failure, secondary to tuberculosis, is reported.' |

Recent work on SemRep has focused on assigning confidence scores capturing the factuality of extracted SemRep triples [110]. These results suggest a promising path for constraining SemMedDB predications further.

#### Causal irrelevance

Machine reading errors have the potential to introduce additional bias. Including inappropriate or irrelevant information can variously (positively or negatively) bias estimates. Irrelevant covariates may be classified into two categories: 1.) variables that are causal relevant (adjacent) but related inappropriately to the exposure and outcome; and 2.) variables that are entirely causally irrelevant. In the first category of irrelevance, individual covariates may be causally relevant but have the wrong type of relevance for the question at hand, as is the case with colliders and mediators mentioned earlier. Controlling for these covariates may result collider bias [111, 106, 112]. An example of collider bias would be controlling for fever when determining a hypothetical relationship between influenza and food poisoning [2]. In the case of mediators, if the intermediate variable’s relationship to the outcome is more vital than that between the exposure of interest and the outcome, then the estimate may be negatively biased.

We were curious to see how including variables selected at random would affect our adjustment and effect estimation procedures. To interrogate the notion of causal relevance and its corollary, causal irrelevance, we have devised, run, and summarized an additional experiment.

We hypothesized that controlling for randomly selected covariates (that were presumed to be irrelevant) would result in more biased models than models incorporating literature-derived confounders. To this end, we built models using the same EHR-derived clinical narrative data and reference dataset. We identified and controlled for up to ten covariates selected at random to empirically evaluate the effect of controlling for irrelevant covariates on error propagation on the outcome, measured by the amount of bias. In the next few paragraphs, we report on our findings.

Figure 6 illustrates the treatment effect estimates from drug / adverse event pairs using different sets of confounders. We ran the procedure over all of the drug / adverse event pairs described earlier. However, to obtain a clearer picture of the methods performed, we removed outliers by excluding models from drug / adverse event pairs scoring in the lowest and highest quartiles. This procedure resulted in slightly different baselines from the entire subset described earlier since there are fewer sample pairs per adverse event.

**Figure 6:**
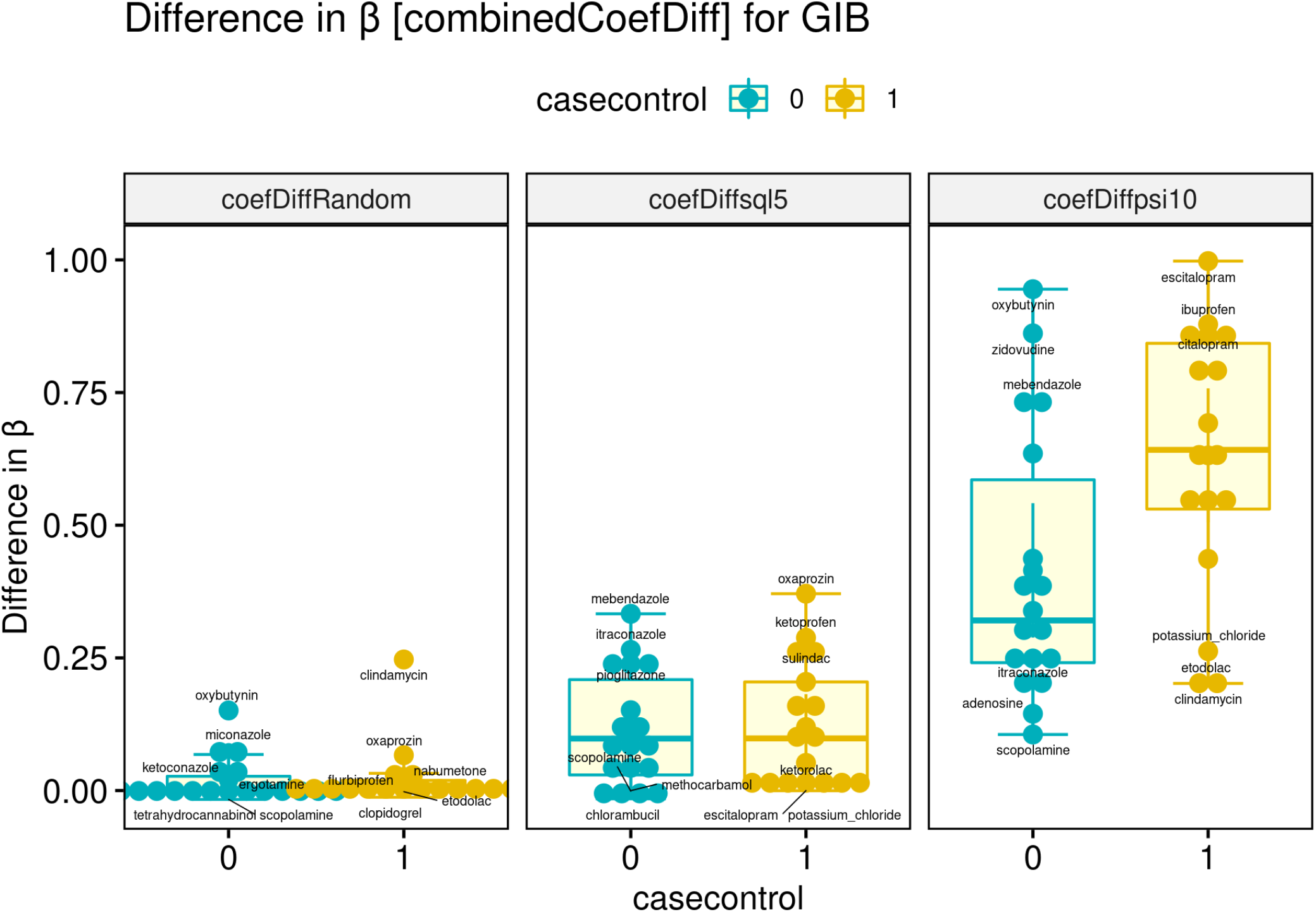
These boxplots represent the differences in unadjusted vs. adjusted regression coefficients (*β*) for gastrointestinal hemorrhage. The leftmost boxplot shows these differences in *β* using randomly selected (irrelevant) covariates whereas the next two boxplots show the differences from using *sql*^5^ and *psi*^10^ confounder search. Controlling for literature-derived methods resulted in more considerable reductions in the negative controls’ estimates than randomly selected covariates. The boxplots generated for from the *β* difference for a drug/adverse event pair sample size of *n* = [17+, 18−]*pairs*. The highest and lowest scoring quartiles were excluded from the counts and statistics reported below.

The leftmost boxplot illustrates the distribution of regression coefficients using ten randomly selected covariates, while the other two boxplots represent regression coefficients from models incorporating literature-derived confounder candidates (*sql*^5^ and *psi*^10^ confounder search).

Of the two literature-derived methods, *psi*^10^ better reduces the negative control pairs’ estimates. The effect estimates for the positive drug / adverse event pairs are slightly higher than the estimates of the negative control pairs on average. The raw regression coefficient scores average treatment estimate scores were not as strongly discriminative.

These observations prompted us to revisit the regression coefficients from the rest of the research data. Table 9 shows the AUROCs from ranking the drug / adverse event pairs by calculating the difference between adjusted vs. unadjusted regression coefficient across different adverse event types. In this way, we were able to perform a direct comparison of the performance of models using randomly selected vs. the literature-derived confounders ((*psi*^10^)).

**Table 9.** This table shows baselines from calculating the AUROCS for the unadjusted *β*s vs. the difference in the unadjusted vs. adjusted regression coefficients from the set of models with randomly selected covariates (middle column with numbers) vs. models with literature-informed confounders in the right column. AKI = acute kidney failure. ALI = acute liver failure. AMI = acute myocardial infarction. GIB = gastrointestinal bleeding.

| AE \ Predictor | Unadj. $\beta$ [Baseline] | $\beta$ Difference [Random] | $\beta$ Difference [$psi^{10}$ confounders] |
| --- | --- | --- | --- |
| AKI [10+, 8-] | 0.762 95% CI: [0.377-1.0] | 0.619 95% CI: [0.156-1.0] | **0.952 95% CI: [0.82-1.0]** |
| ALI [33+, 11-] | 0.587 95% CI: [0.363-0.811] | 0.636 95% CI: [0.436-0.837] | **0.656 95% CI: [0.482-0.837]** |
| AMI [9+, 15-] | 0.667 95% CI: [0.412-0.922] | 0.541 95% CI: [0.290-0.792] | 0.563 95% CI: [0.311-0.815] |
| GIB [17+, 18-] | 0.680 95% CI: [0.495-0.865] | 0.533 95% CI: [0.332-0.733] | **0.722 95% CI: [0.545-0.90]** |
| Overall | 0.561 95% CI: [0.452-0.669] | 0.541 95% CI: [0.429-0.653] | **0.662 95% CI: [0.562-0.763]** |

#### Treatment-confounder feedback

We made simplifying assumptions that assumed that confounder behavior would be stable for the practical purpose of reducing confounding bias. However, it is known that the relationship between biomedical entities can vary with time. Such is the case, for example, when a therapy affects the indication for which it is prescribed that is itself an independent cause or risk factor of the disease. In this setting, a confounder variable also behaves like an intermediate variable or mediator. Mediators are links on the causal chain between exposure and outcome: *A*_*exposure*_ → **mediator** → *Y*_*outcome*_.

The modeling problem associated with the issue of such problematic covariates is called treatment-confounder feedback [113, 114, 115]. Treatment-confounder feedback can result in “overcontrol bias,” wherein estimates are biased toward the null hypothesis [95]. Bias towards the null hypothesis occurs if the intermediate variable’s effect overwhelms the direct effect from the exposure. For reference literature on this phenomenon, see chapter 20 in [116].

For instance, literature search ranks diabetes as a probable confounder of the anti-diabetic drug rosiglitazone and acute myocardial infarction. If rosiglitazone is effective at treating diabetes, the effect of treating diabetes might reduce the risk of heart attack. If diabetes is a more powerful risk factor than rosiglitazone, controlling for diabetes without truncating the data longitudinally may bias conclusions. In this study, the case of acute kidney failure presented an overall pattern of overcontrol bias from treatment-confounder feedback in which the *β* scores from positive cases were lower on average than the adjusted scores in the negative controls, after incorporating the literature-derived confounders, violating the assumptions mentioned earlier that the effect estimates of the negative controls should tend towards zero (ASSUMPTION I) and the estimates of the positive controls should be greater than the negative controls (ASSUMPTION II).

Since knowledge is incomplete and imperfect, misspecified models, or models that erroneously include or omit certain variables, are likely the norm in causal modeling. It is also likely that treatment-confounder feedback introduces additional hurdles to bias reduction. Fortunately, the question of how to handle confounding in these circumstances has motivated the developments causal parameter estimation methods methods that can address treatment-confounder feedback [117]. Advanced estimation frameworks such as that of targeted learning are robust to model misspecification by employing data-adaptive procedures that combine a propensity score model with an outcome regression to optimize causal effect estimates, which can be further enhanced with ensemble machine learning [118, 119]. For more details, a paper is available here [120] outlining how to apply targeted learning procedures that also offers important insights concordant with the observations made in the present study.

### 5.4. Limitations and themes for future work

Ideally, we would wish that our methods for studying observational data would possess sufficient rigor to approach the level of scientific confidence of an RCT. We have enumerated additional limitations of the present study with a view toward future work in this area to bridge gaps in the following areas:

- **IMPROVING DATA**
  – **Improving data hygiene:** We did not carry out advanced phenotyping procedures, correct for missingness in the data, or rigorously define exposure and outcome criteria. Although we have no evidence to prove that such factors negatively impacted our methods’ performance, these factors likely set a ceiling on the performance improvements attainable by applying our methods. In our current work, we have adopted more rigorous approaches to measure and improve the validity of the clinical data [121, 122]. Other improvements could be attained with more extensive vocabulary mapping. For example, we have not reconciled the data underlying each synonym into a single representation for each overarching biomedical entity of interest representing confounder candidate concepts. In future work, we may explore NER systems beyond or additional to MedLEE, e.g., using ensemble-based NER methods [78], and take efforts to approximate a target trial [123] by better defining the observation periods and setting “time zero.”
  – **Addressing temporality:** one fundamental limitation of our approach stems from the coarseness of cross-sectional data. Cross-sectional data represent a “snapshot in time” rather than temporality. These data may violate independence and exchangeability assumptions [124] and thereby introduce “immortal time bias” [123]. Study designs that rigorously apply eligibility criteria explicitly specifying a “time-zero” and observation periods for the exposure, outcome, and confounding variables can address this limitation. Longitudinal data may also be important for optimal performance.
  – **Structured + unstructured data:** In the present study, we used empirical data extracted from the unstructured free-text narrative. EHR systems were designed primarily for administrative record-keeping such as claims, billing, and other non-research-specific information and to fulfill legislative mandates [125]. Anticipating administrative needs, system designers naturally used structured fields to capture the most critical information for these purposes. However, unstructured text can provide unique insight to inform both clinical ends and basic science [126]. For example, information that may be absent in structured fields (as was the case in [79]) about sub-pathological states such as worry, hypomania, or other affective states captured by clinicians and present in clinical notes could provide insight into the progression of chronic illnesses outcomes such as Alzheimer’s disease and other neurodegenerative diseases. Many other types of valuable information may be missing in structured data, including the reasons for drug discontinuity events [127]. Thus unstructured data is necessary to enhance our understanding of risk factors as well as potential benefits from drug exposures. For a more complete picture of patient health, it will be essential to capture the valuable information from both structured fields and unstructured free-text in the EHR systems in future work. However, to take full advantage of the richness of free-text clinical data in EHR systems, additional efforts are needed to mitigate data quality issues.
- **IMPROVING KNOWLEDGE**
  – **Confounder quality control:** Machine reading produces errors, and those errors can propagate additional biases. As we saw in the randomly selected variable experiment, controlling for etiologically irrelevant randomly selected covariates did not help improve the detection of drug / adverse event pairs. There is no way of knowing whether the measures of association represent artifacts of the data sample rather than support for a causal effect. On the other hand, machine reading errors may also introduce a variable that is causally related to the exposure and outcome of interest in undesirable ways. For example, if machine reading errors introduce a collider into an adjustment set, then collider or selection bias can occur, amplifying bias.
  – **Proxy confounders:** While automated tools such as those described in this paper may be useful, these tools are only useful for identifying confounders from *a priori* knowledge in the published data, and residual bias may remain from uncontrolled confounders. Machines still have a very limited ability to understand language and causality. It is crucial that researchers familiarize themselves both with confounders from the literature, the features of their data, and engage with subject-matter experts familiar with the local causal structure of biomedical entities involved [120]. Seemingly irrelevant covariates may still be useful for capturing missing information, even though they are not etiologically related to either the exposure or the outcome directly. For example, information about whether patients play ice hockey may seem irrelevant on the surface. However, such information could be potentially useful for controlling for the effect of socioeconomic status since hockey is an expensive sport. Thus, detailed knowledge of local causal structures is crucial for formulating a working model that can most reliably help test causal hypotheses from observational data and covariate selection [32]. However, such procedural knowledge is likely beyond the capability of any AI system for the conceivable future. For these methods to be of use in settings where significant precision is required, it is necessary to have a human possessing substantial subject-matter knowledge in the loop.
  – **Using improved knowledge representation:** Our methods may benefit from updated literature-based discovery and distributional representation methods. For example, Embedding of Semantic Predications (ESP) is a neural-probabilistic extension of the PSI model that has been shown to improve performance in predictive modeling tasks [128]. Still, other comparator methods exist, which we will also soon explore [129].
  – **Including other discovery patterns:** It is probably not the case that a single discovery pattern is sufficient to capture all confounders. More research is needed in this area to expand search through other pathways.
  – **Identifying ‘minimal adjustment sets’:** Although the thresholds on the number of confounder candidates incorporated into the models are to some extent justified from a heuristic standpoint on the basis of examining the number of confounders per drug / adverse event pair, we note the arbitrary nature of these thresholds as a limitation. However, the selection of optimal subsets of confounders is a distinct and active research area referred to as causal feature selection [130], and further optimizing feature selection for causal inference is not the focus of this paper. We shall seek to remedy this shortcoming by applying minimal adjustment set methods in future work.
  – **Developments in machine reading:** Limitations in our capability to process knowledge into a usable form further limit our ability to control for confounding. The SemRep system is currently being upgraded to support end-user extensibility and with exciting features that include methods for encoding factuality levels [110], where factuality levels denote the degree of belief in mined assertions from text. Factuality levels are potentially useful for improving “knowledge hygiene” and for improving the detection and identification of contradictory claims in the literature [131]). The Integrated Network and Dynamical Reasoning Assembler, or INDRA, NLP system can translate scientific prose directly into executable graphical models [132, 133]. INDRA output can be filtered through additional postprocessing filters to provide confidence scores [134]. Such additional information weighting the validity of machine reading output could help to screen out irrelevant literature-derived covariates. To this end, we (authors S.A.M. and R.D.B. along with others) are exploring combining knowledge from the SemRep reading system with knowledge extracted using the INDRA system [132] into an ontology-based knowledge graph platform populated with comprehensive biomedical knowledge [135].
  – **The deconfounder:** The extent of knowledge itself further limits the ability to control for confounding (and also the observation that the claims of most scientific studies are invalid [136]). There have been notable efforts afoot to contrive a synthetic variable called a deconfounder [37, 137]. Deconfounders can substitute for substantive knowledge of the causal structure relative to the exposure and the outcome and also potentially correct for missingness. An intriguing line of study, as yet unpursued, would be interesting to combine both simulated and empirically-derived confounders.
- **IMPROVING ESTIMATION**
  – **Advanced estimation methods:** More advanced effect estimation techniques exist (e.g., G-methods [115, 114, 138, 139], Inverse Probability Weighting [140], and targeted learning estimators [118]) with more desirable statistical properties than the method presented here. These tools are more resilient against issues that commonly plague observational studies including model specification errors from treatment-confounder feedback and missing-not-at-random data. Accordingly, these more advanced estimation methods are a natural fit with the knowledge integration methods outlined in this paper (see [120] for an overview).
  – **More appropriate target causal parameter:** Also, causal effect parameters such as Effect of the Treatment on the Treated (ETT) are often more appropriate for situations where treated subgroups may have distinct background characteristics compared with untreated subgroups, as is often the case with observational data.

### 5.5. Conclusions

This paper introduced a generalizable framework for helping solve the ubiquitous problem of confounding bias, an endemic threat to all observational studies, and expands upon our previous work combining computable knowledge from the literature with observational data to reduce confounding [26, 70].

We have shown that our knowledge integration methods can modestly improve the ability to detect genuine pharmacovigilance signals from observational clinical data, though leaving ample room for methodological refinement, given the limitations listed above.

Notwithstanding confounding adjustment, residual bias may remain from unmeasured, mismeasured, or omitted variables [96], as well as from other forms of systematic bias [141] along with random noise. However, we assume that confounding and other forms of bias can be reduced but not eliminated.

We found that incorporating literature-derived confounders more often than not improved causal inference using a publicly available reference dataset and that confounder candidates from semantic vector-based search improved the detection of genuine drug safety signals over string-based confounder search.

Moreover, semantic vector-based search offered improved performance over string-based search. We conjectured that the advantageous properties of the representation scheme helped prioritize the most useful confounder candidates and resulted in models with the most improvement over baseline. Our framework can be easily adapted to help in other areas, such as estimation of therapeutic rather than adverse effects, with only a modicum of difficulty. This provides an avenue through which to leveraging existing causal knowledge to to accelerate the discovery of new knowledge, by improving the reliability of causal inference from observational data.

## Data Availability

Following IRB approval and a data usage agreement, we obtained permission to use the EHR data for this study. These data require approval to access. However, we have included the models and other analytic data from this project.

https://github.com/kingfish777/causalSemantics

## Acknowledgments

This research was supported by the US National Library of Medicine grants: R01 LM011563, 2 T15 LM007093−26 (a training fellowship from the Keck Center for Interdisciplinary Bioscience Training of the Gulf Coast Consortia), 5 T15 LM007059−32, NIH/BD2K supplement R01 LM011563−02S1, NCATS Grant U54 TR002804, Cancer Prevention Research Institute of Texas (CPRIT) Precision Oncology Decision Support Core RP150535, and CPRIT Data Science and Informatics Core for Cancer Research RP170668. Many thanks to readers Doug Landslittel, Harry Hochheiser, the causal reading group at Carnegie Mellon University, and to the anonymous peer-reviewers for helpful suggestions on earlier drafts of this manuscript. The opinions expressed in this manuscript do not necessarily reflect those of the National Institutes of Health or the National Library of Medicine.

## 6. CRediT authorship contribution statement

Conceptualization, S.A.M., T.C., E.V.B., and P.W.; Methodology, S.A.M. and T.C.; Software, T.C. and S.A.M.; Formal Analysis, S.A.M.; Investigation, S.A.M.; Resources, T.C., E.V.B., R.D.B.; Data Curation, S.A.M.; Visualization, S.A.M.; Writing - Original Draft, S.A.M.; Writing - Review & Editing, S.A.M., T.C., R.D.B., E.V.B., P.W.; Funding Acquisition, T.C., E.V.B., R.D.B.; Supervision, T.C. and R.D.B.

## A. Appendix A - Stopwords

### A.1. Stopterms

*The following terms were excluded from being confounder candidates on account of the terminological vagueness or other reasons*. adhesions; adolescent; adult; agent; animals; antibodies; antigens; apoptosis; application procedure; assay; assessment procedure; assessment procedure; bacteria; biopsy; blood; body tissue; boys; canis familiaris; capsule; cattle; cell line; cell membrane; cells; cerebrovascular accident; child; chronic disease; clinical research; cohort; color; complication; congenital abnormality; contrast media; control groups; country; detection; diagnosis; disease; dna; elderly; embryo; entire hippocampus; enzymes; excision; extracellular; family; family suidae; felis catus; fibroblasts; follow-up; fracture; fume; functional disorder; genes; girls; growth; house mice; human; implantation procedure; implantation procedure; individual; induction; infant; infiltration; injection procedure; injection procedure; injury; intervention regimes; jersey cattle; lesion; macaca mulatta; macrophage; magnetic resonance imaging; male population group; malignant neoplasm of breast; malignant neoplasms; management procedure; management procedure; medical imaging; membrane; micrornas; mild adverse event; monkeys; monoclonal antibodies; mothers; mus; muscle; nonhuman primates; obstruction; operative surgical procedures; organ; participant; pathogenesis; patient; patient state; patients; persons; pharmaceutical preparations; pharmacophore; pharmacotherapy; placebos; plants; plasma; primates; procedures; prophylactic treatment; proteins; protoplasm; psychopharmacologic agent; rabbits; radiation therapy; rats; rats; rattus norvegicus; receptor; rna; rodent; screening procedure; screening procedure; screw; serum; sloths; solutions; stimulation procedure; stimulation procedure; study models; substance; supplementation; symptoms; syndrome; techniques; test result; therapeutic procedure; toxic effect; transplantation; treatment aids; treatment protocols; voluntary workers; water; woman; young child

